# Clinical prediction with localized modeling using similarity-based cohorts: A scoping review

**DOI:** 10.1101/2024.06.04.24308433

**Authors:** Adi Cohen, Patti McCall-Junkin, Jason Cory Brunson

**Affiliations:** College of Osteopathic Medicine, Nova Southeastern University Davie, FL; Smathers Libraries Academic Research & Consulting Services, University of Florida Gainesville, FL; Laboratory for Systems Medicine, University of Florida Gainesville, FL

**Keywords:** case-based reasoning, localized modeling, nearest neighbors, clinical decision support, scoping review

## Abstract

**Background:** Applications of classical case-based reasoning (CBR) have given rise to a family of techniques we call “localized models”, in which a statistical model is fitted to a neighborhood of labeled cases matched by similarity to a target case. We aim to describe clinical and health applications of localized models to date and propose a general framework for their design and evaluation.

**Methods:** We searched four bibliographic platforms during 2021 July 19–22, updated 2024 January 24. We set four eligibility criteria to identify applications of localized models to clinical and health tasks. Two authors divided title/abstract screening and reviewed screened entries for inclusion. We discussed settings, tasks, and tools; identified and tabulated themes; and synthesized the methods into a general framework.

**Results:** Of 1,657 search results, 360 were reviewed, then combined with 43 publications that seeded the review and 1 obtained by citation tracking. 27 were included, published 1997–2022. The specificity of search terms was poor, and inter-rater reliability was low. Almost all models were predictive, the most common tasks being prognosis and diagnosis. Most studies used clinical, occasionally laboratory and image, data. Several addressed memory and runtime costs. A general technique that specializes to most of those reviewed involved matching, retrieval, fitting, and evaluation steps that could optionally be supervised, optimized, or recursively performed.

**Conclusions:** Localized models have potential to improve the performance of clinical decision support tools while maintaining interpretability, but rigorous comparisons to competing methods must be conducted and computational hurdles must be overcome. We hope that our review will spur future work on efficiency, reproducibility, and user needs.

## 1 Introduction

A main driver of the development of clinical information systems (CIS) and clinical decision support (CDS) tools has been to leverage the advantages of routinely collected health data toward improving care. These advantages include their immediate accessibility through the institutional EHR, billing database, or other source; their specificity to the institution and the population it serves; and the closeness in time of the available data. Though by definition not collected for research use, the secondary research use of routinely collected health data has been put forth, as “practice-based evidence”, a complement to the paradigm of evidence-based practice.

Over the same period of time, interest in individualizing care from population-derived evidence-based recommendations to specific patient needs has driven the application of advanced computational tools, including artificial intelligence (AI). Whereas classical models produced formulae involving a limited set of data elements that would be uniformly applied to all patients, AI models often process hundreds or more variables in opaque ways to yield predictions that depend on not only their values but their associations and interplay with each other. The loss of straightforward interpretations of these models has limited their practical uptake and motivated the construction of explanatory statistics for opaque models as well as the development of more interpretable complex models.

One of few methodologies to address all three of these established needs is one we call *localized modeling*, which appears to have been introduced several times independently under different names, and in slightly different forms. The approach is a specialized form of case-based reasoning (CBR) that relies on a patient similarity measure^1^ to extract a cohort of past or training cases that then inform the diagnosis, prognosis, or care of a new or test case. Traditional CBR returns these retrieved cases to inform human decision-making; the related nearest neighbors (NN) technique generates predictions from cohorts automatically via averaging (regression) or voting (classification) of their outcomes. In contrast, localized modeling fits families of predictive models, for example generalized linear models (GLMs), to retrieved similarity cohorts in order to generate predictions. Localized models thus provide an individualized way to fit fully interpretable models to large population data.

This approach harkens to the aspirational “green button” that would, in response to a query, automatically retrieve patient data from an institution’s records with which to conduct on-demand retrospective studies for an individual patient (Longhurst, Harrington, and Shah 2014). Several hurdles face the deployment of such an approach in practice, but its benefits must first be demonstrated. Our motivations in this scoping review are to describe the settings in and problems with which this approach has been tasked, to attempt to organize them within a common framework, and to evaluate the promise they show toward achieving this or other needs in clinical informatics. Along the way, we attempt to reconcile terminology, summarize motivations and evaluations, and propose valuable follow-up work.

### 1.1 Related work

Clinical CBR emerged among rule-based approaches and other AI tools in the development of expert systems (Aamodt and Plaza 1994). Early implementations realized a general workflow described as “the four REs”, later the “R4 cycle” (Aamodt and Plaza 1994; Begum et al. 2011): Given a new case (or problem), the system *retrieves* one or more past cases (solved problems) from a corpus, *reuses* these to generate an understanding of (solution to) the new case, *revises* this understanding (solution) to better fit the new case (sometimes called “adaptation”), and *retains* the new case and its eventual understanding (solution) in the corpus to be retrieved and reused in future. In contrast to rule-based systems, which are variable-based and generally interpretable as a single rule applied to all new cases, case-based systems provide case-specific interpretations in the form of a number of more fully understood reference cases. While the similarity measure used in the retrieval step need not be changed as the corpus grows, proposed measures have been diverse, contested, and rarely systematically compared.

Kolodner (1992) distinguished two styles of CBR: problem-solving, which is more procedural and used when objectives are more clearly defined, and interpretative, which provides categorizations and justifications for possible solutions. A similar dichotomy is commonly used to distinguish the performance criterion for the usefulness of predictive models from the interpretability criterion for their usability. In these terms, localized models are highly procedural: The step of fitting a predictive model to a retrieved cohort is an automated adaptive strategy (Begum et al. 2011), and the parameters that govern cohort retrieval can be tuned alongside model hyperparameters in a machine learning (ML) workflow. Accordingly, localized models have been primarily used for and evaluated on their ability to predict classes or outcomes. Nevertheless, as some recent applications have shown, they can be used to draw inferences about the importance of different risk factors to specific individuals. While most ML models come equipped with measures of feature importance and model-agnostic tools can generate explanations for model predictions, these quantifications are not directly interpretable model components analogous to the split nodes of a decision tree (DT) or the coefficients of a GLM. While the patient similarity measure used to retrieve each cohort may be complicated, the cohort itself can be directly inspected by the user. Provided the model family fitted to the cohorts is interpretable, the localized model inherits this property. Thus, localized models may embody both styles of CBR.

We refer the interested reader to several previous reviews of CBR in medicine (Gierl, Bull, and Schmidt 1998; Begum et al. 2011; Choudhury and Begum 2016), of measures of patient similarity (Dai, Zhu, and Liu 2020), and of uses of patient similarity in predictive models (Welch and Kawamoto 2013; Sharafoddini, Dubin, and Lee 2017; Parimbelli et al. 2018). While the reviews of CBR focus on applications using health data, the patient similarity reviews encompass many additional types of data (various molecular -omics, genetic tests, medical images, laboratory tests, patient preferences, patient-reported outcomes, tracking devices, social media) and survey a much broader scope of models (exploratory analysis via dimension reduction and cluster analysis; risk evaluation and outcome prediction; clinical decision support and software tools). For the present review, we are interested in how similarity matching on patient-level health data is used to construct local cohorts for predictive or inferential modeling.

### 1.2 Objectives

Our goals in this review are (1) to describe applications of similarity-based localized models using patient-level health data and (2) to provide a general framework for the design and evaluation of such localized models. We focus narrowly on localized models, rather than broadly on patient similarity–based clinical decision support tools, so that we may thoroughly assess their value in terms of reported evaluations and comparisons to other methods. This also allows us to devise a high-level framework that specializes to the majority of these methods, which we then use to frame our discussion and recommendations.

Our selection of relevant papers from the search corpus will be based on the following inclusion/exclusion criteria:

- Uses case-level data from a corpus of past cases with known responses (response may be outcome, diagnosis, subtype, etc.)
- Defines a numeric multivariate case similarity measure (allow integer-valued measures)
- Uses the similarity measure to retrieve cohorts for index cases from the corpus (for example, based on a training–testing partition)
- Fits statistical models to cohorts from which to make predictions or draw inferences about index cases

(outcome predictions, survival estimates, risk factor contributions, model evaluation statistics, etc.)

We expected the general framework to come down to three choices: A patient similarity measure, a cohort selection process, and a statistical model family. A general implementation based on this method would allow researchers to expedite every step of the analysis process, including retrieval, optimization, and evaluation, and enable sensitivity, robustness, and multiverse analyses that help identify the most consequential choices along the way.

## 2 Methods

Here we describe our review process, including deviations from plans and the reasons for them. More details are included in Section 4.6.

### 2.1 PRISMA checklist

We include a PRISMA checklist as Supplemental Table 1 and a PRISMA abstract checklist as Supplemental Table 2. Because we focus on methodologies rather than conceptual approaches or evidence, we deviate in some ways from PRISMA guidelines. In particular, those items of the checklist involving bias assessment and quantitative synthesis are intended for meta-analyses so did not apply to this study. Because we are not aware of any standard procedures for conducting reviews and syntheses of methodology, no protocol was prepared for this study.

### 2.2 Search

We derived the eligibility criteria itemized in the Introduction from a seed set of previously read studies. Based on these criteria, we formulated search strings for five platforms: PubMed, Web of Science, Academic Search Premier, Google Scholar, and MathSciNet. We finalized the PubMed search first, then adapted it to the other platforms (Section 4.6).

Through each platform, we searched those databases included by default. This means that we searched both MEDLINE and PubMed Central (PMC) through Pubmed (we did not exclude other databases, but our earliest included results post-date them) and that we searched the six indices of the Core Collection (the Science Citation Index Expanded, the Social Sciences Citation Index, the Arts & Humanities Citation Index, the Emerging Sources Citation Index, the Conference Proceedings Citation Index, and the Book Citation Index) as well as several regional databases through the Web of Science platform.

The structure and terms of our search strings were inspired in part by previous reviews adjacent to our topic of interest (Sharafoddini, Dubin, and Lee 2017; Parimbelli et al. 2018). We organized the PubMed search in disjunctive normal form (an OR of ANDs). Following the solidification of an outline, we expanded each term to include synonyms that are similar enough to be applicable to our search. We then evaluated the expanded search string using the PubMed Advanced Search platform. We initially included each term and their synonyms separately to evaluate what resulted. We pruned several terms that returned no results (“phrases not found”) or to reduce the number of results. At the conclusions of this process for each individual term, we combined the search terms using disjunctive normal form.

We conducted all searches over 2021 July 19–22. We tentatively excluded results from Google Scholar because they were missing abstracts, and later agreed to discard these results due to the lack of reproducibility of the search. We organized the remaining results into a public Zotero collection with one subcollection for each database. We then imported the pooled results to Covidence, which merged some duplicate entries.

We repeated the search on 2024 January 24 on the PubMed and Web of Science databases to bring the results up to date.

### 2.3 Title/abstract screen

Within Covidence, we screened titles and abstracts for relevance. We decided on four eligibility criteria to expedite the screening process. Articles must be written in English, for readability; they must be original studies, to exclude secondary sources with duplicate information; their use settings must be medical, clinical, or related, to keep our review topical; and it must not be clear that their use of our search terms was different from our intended meaning. Each of two authors (AC and JCB) screened roughly half of the entries. They regularly reviewed each other’s decisions to improve consistency. In cases of uncertainty, entries were included. For the update, one author (PM) screened all results for three criteria and another (JCB) screened the survivors for the fourth criterion.

### 2.4 Full text review

We set out 4 criteria for full-text review to restrict to studies that used some form of localized modeling on health data:

- Pulls case-level data from a corpus of past cases with known classes or outcomes
- Defines a numeric multivariate case similarity measure
- Uses the similarity measure to select cohorts for index cases from the corpus
- Fits statistical models to cohorts to make predictions or inferences about index cases

Note that classical CBR satisfies the first and third criteria by definition and in most cases will satisfy the second.

In Covidence, two authors (AC and JCB) independently evaluated each manuscript for these eligibility criteria. The first criterion that a manuscript failed was designated the reason for exclusion. An antecedent criterion was used to exclude manuscripts that did not report the results of original studies involving real-world experiments or empirical data, for example surveys of prior work and proposals of frameworks. In cases of disagreement between the authors on the reason for exclusion, the first criterion was adopted. The authors arrived at agreement on inclusion or exclusion through discussion. We calculated inter-rater reliability to evaluate our screening and review process.

During full text review, we decided to expand the conception of statistical models (fourth criterion): Rather than restricting to models that are fit and evaluated in separate steps, we chose to allow simple statistical summaries such as mean survival (Mariuzzi et al. 1997) and survival curves (Lowsky et al. 2013). These approaches were novel to CBR and presage later developments, so were helpful to understanding the development of localized modeling. However, this then admitted studies that applied conventional nearest neighbors prediction: Each retrieved cohort consisted of the *k* most similar cases to the index case, and the response for the index case was predicted to be either the mean (continuous response) or the plurality (discrete response) of the cohort’s responses. A review of all studies that use nearest neighbors prediction would be impractical. Because our focus is on novel approaches that combine similarity-based retrieval and statistical modeling of retrieved cohorts, we chose to exclude those studies whose approach was equivalent to nearest neighbors prediction using a conventional similarity measure.

Finally, one author (JCB) applied the same review process to the seed set of 43 studies that inspired the review. The same author later reviewed the updated results.

### 2.5 Coding

We next collected characteristics of included studies. The features included bibliographic fields (date of publication, journal, authors, title, keywords, DOI), study goals (objective, generalizable knowledge, evaluation, clinical/medical domain), data sets (data source, type of data, range of data, number of cases/incidences, number of predictors/features), and methodological choices (types of similarity measure, families of adaptation step/statistical model, method(s) compared against, performance measures, results of evaluations and comparisons, name given to modeling approach). We used these data to detect and visualize trends amongst the included studies.

### 2.6 Synthesis

Rather than an evidence synthesis characteristic of most systematic reviews, we here pursue a methodology synthesis to harmonize largely independent research efforts that have converged on a common family of techniques. The goal will be to describe a unified framework for localized models that can be used to guide future study designs and implementations as well as more systematically evaluate variations on the theme and measure the dependence of results on modeling choices.

## 3 Results

### 3.1 Selection

Figure 1 depicts our identification of studies via databases and registers. Following the completion and input of each search string, there were a total of 1,817 sources within all of the platforms used. De-duplication resulted in 1,657 entries, which were added to the title/abstract screening for review. Of these, 360 entries met the screening criteria and were assessed through full text review. Of these, 51 fit the original criteria, and 21 were included as distinct from NN prediction.

**Figure 1:**
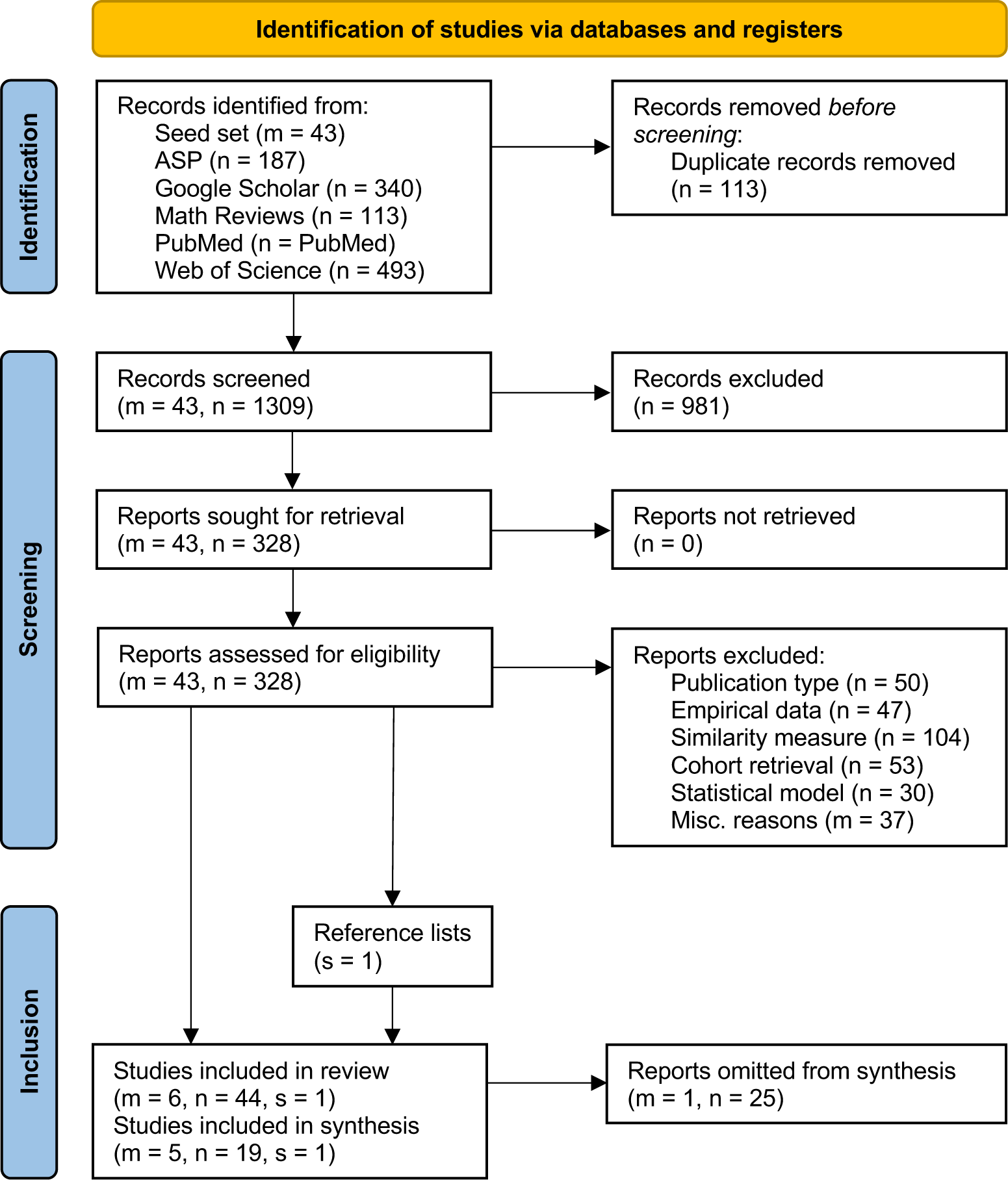
PRISMA-S flow chart. Lowercase letters refer to items obtained from the seed set (m), the structured search (n), and citation/reference tracking (s). Within the structured search results, summands correspond to original and update.

From the first search results, there were 60 disagreements over inclusion versus exclusion. Inter-rater reliability was low, at 82% relative to a 72% probability of random agreement. Only one author was available to review full texts from the update search.

We then reviewed 43 studies comprising a seed set that inspired this review. After removing duplicates and screening for eligibility, we were left with 6 additional studies (Park, Kim, and Chun 2006; Lowsky et al. 2013; Lee, Maslove, and Dubin 2015; Ng et al. 2015; Lee 2017; N. Wang et al. 2019), 1 of which was excluded from the synthesis for using NN prediction. Reference tracking from the 43 + 360 = 403 studies assessed for eligibility led us to identify 1 additional study that met our criteria (Kasabov and Hu 2010). This left us with 21 + 1 + 5 = 27 studies included in the review and synthesis.

### 3.2 Bibliographic and methodological properties

The 27 included studies were analyzed based on several characteristics, and we report and describe some observations here (Table 1). The studies were published in a variety of journals, with some of greater frequency, though no single journal published more than 3. The journal *Evolving Systems* published 2 of the included studies, *Artificial Intelligence in Medicine* published 3, *Hindawi - Journal of Healthcare Engineering* published 3, *Evolving Systems* published 2, and *PLOS One* published 2.

**Table 1:**
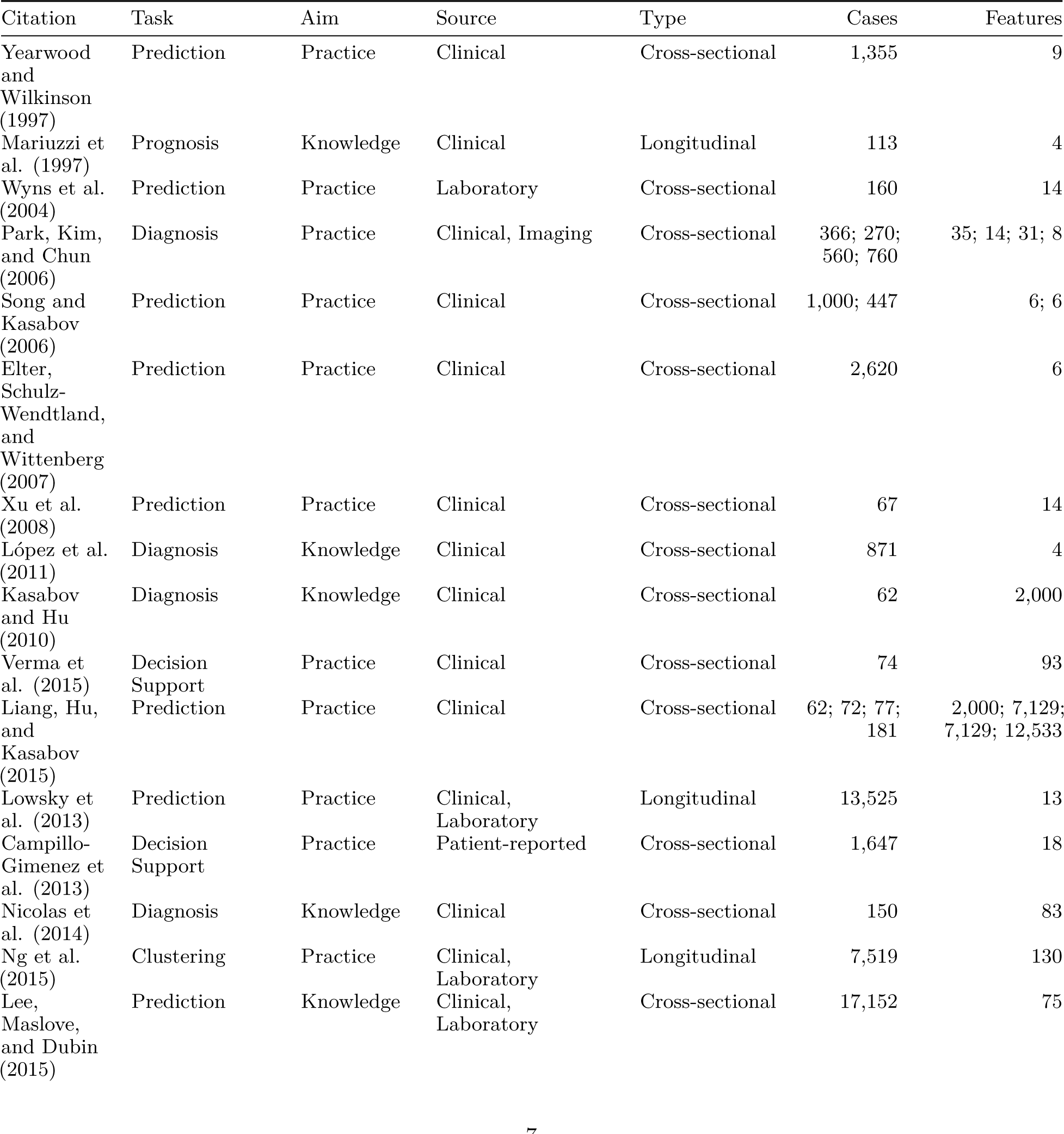
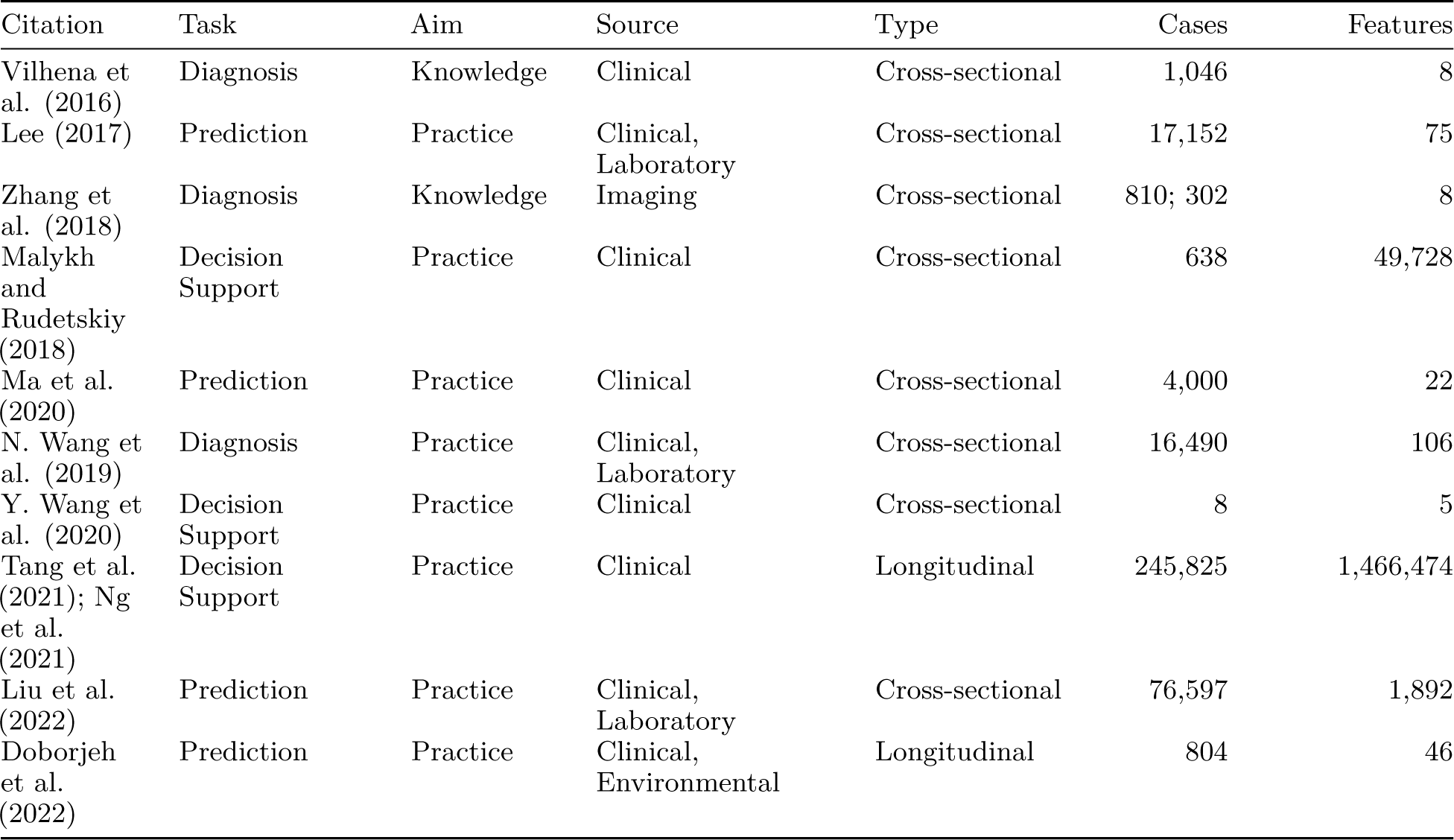
Studies included in the method synthesis, arranged by the earliest the study is known to have been public.

| Citation | Task | Aim | Source | Type | Cases | Features |
| --- | --- | --- | --- | --- | --- | --- |
| Yearwood and Wilkinson (1997) | Prediction | Practice | Clinical | Cross-sectional | 1,355 | 9 |
| Mariuzzi et al. (1997) | Prognosis | Knowledge | Clinical | Longitudinal | 113 | 4 |
| Wyns et al. (2004) | Prediction | Practice | Laboratory | Cross-sectional | 160 | 14 |
| Park, Kim, and Chun (2006) | Diagnosis | Practice | Clinical, Imaging | Cross-sectional | 366; 270; 560; 760 | 35; 14; 31; 8 |
| Song and Kasabov (2006) | Prediction | Practice | Clinical | Cross-sectional | 1,000; 447 | 6; 6 |
| Elter, Schulz-Wendtland, and Wittenberg (2007) | Prediction | Practice | Clinical | Cross-sectional | 2,620 | 6 |
| Xu et al. (2008) | Prediction | Practice | Clinical | Cross-sectional | 67 | 14 |
| López et al. (2011) | Diagnosis | Knowledge | Clinical | Cross-sectional | 871 | 4 |
| Kasabov and Hu (2010) | Diagnosis | Knowledge | Clinical | Cross-sectional | 62 | 2,000 |
| Verma et al. (2015) | Decision Support | Practice | Clinical | Cross-sectional | 74 | 93 |
| Liang, Hu, and Kasabov (2015) | Prediction | Practice | Clinical | Cross-sectional | 62; 72; 77; 181 | 2,000; 7,129; 7,129; 12,533 |
| Lowsky et al. (2013) | Prediction | Practice | Clinical, Laboratory | Longitudinal | 13,525 | 13 |
| Campillo-Gimenez et al. (2013) | Decision Support | Practice | Patient-reported | Cross-sectional | 1,647 | 18 |
| Nicolas et al. (2014) | Diagnosis | Knowledge | Clinical | Cross-sectional | 150 | 83 |
| Ng et al. (2015) | Clustering | Practice | Clinical, Laboratory | Longitudinal | 7,519 | 130 |
| Lee, Maslove, and Dubin (2015) | Prediction | Knowledge | Clinical, Laboratory | Cross-sectional | 17,152 | 75 |
| Vilhena et al. (2016) | Diagnosis | Knowledge | Clinical | Cross-sectional | 1,046 | 8 |
| Lee (2017) | Prediction | Practice | Clinical, Laboratory | Cross-sectional | 17,152 | 75 |
| Zhang et al. (2018) | Diagnosis | Knowledge | Imaging | Cross-sectional | 810; 302 | 8 |
| Malykh and Rudetskiy (2018) | Decision Support | Practice | Clinical | Cross-sectional | 638 | 49,728 |
| Ma et al. (2020) | Prediction | Practice | Clinical | Cross-sectional | 4,000 | 22 |
| N. Wang et al. (2019) | Diagnosis | Practice | Clinical, Laboratory | Cross-sectional | 16,490 | 106 |
| Y. Wang et al. (2020) | Decision Support | Practice | Clinical | Cross-sectional | 8 | 5 |
| Tang et al. (2021); Ng et al. (2021) | Decision Support | Practice | Clinical | Longitudinal | 245,825 | 1,466,474 |
| Liu et al. (2022) | Prediction | Practice | Clinical, Laboratory | Cross-sectional | 76,597 | 1,892 |
| Doborjeh et al. (2022) | Prediction | Practice | Clinical, Environmental | Longitudinal | 804 | 46 |

**Table 2:** Specializations of general framework to included studies. Flags: R = recurse, S = supervised, T = tuned (optimized).

| Citation | Relevance | Retrieval | Adaptation |
| --- | --- | --- | --- |
| Mariuzzi et al. (1997) | Levenshtein distance | Statistical | Survival (S) |
| Yearwood and Wilkinson (1997) | Weighted sum (S) | Combinatorial | Proportion (S) |
| Wyns et al. (2004) | Kohonen mapping | Adaptive | Representation (S) |
| Park, Kim, and Chun (2006) | Euclidean distance | Statistical (T) | Weighted sum (S) |
| Elter, Schulz-Wendtland, and Wittenberg (2007) | Entropic distance | Combinatorial | Proportion (S) |
| Xu et al. (2008) | Euclidean distance | Combinatorial | Proportion (S) |
| Song and Kasabov (2006); Kasabov and Hu (2010); Liang, Hu, and Kasabov (2015); Verma et al. (2015) | Feature selection (R,S) | Combinatorial (R) | Fuzzy rules (S,T) |
| López et al. (2011) | User-defined | User-defined | User-defined |
| Lowsky et al. (2013) | Mahalanobis distance (S) | Combinatorial (T) | Proportional hazards (S) |
| Campillo-Gimenez et al. (2013) | Weighted overlap | Combinatorial (T) | Weighted sum (S) |
| Nicolas et al. (2014) | Distance metric learning (S) | Combinatorial | Association rules (S,T) |
| Ng et al. (2015) | Locally supervised metric learning (S); Feature selection | Combinatorial | Logistic model (S) |
| Lee, Maslove, and Dubin (2015) | Cosine similarity | Combinatorial (T) | Logistic model (S); Decision tree (S) |
| Vilhena et al. (2016) | Range overlap | Clustering (S) | Artificial neural network (S) |
| Lee (2017) | Cosine similarity; Random forest proximity (S) | Combinatorial (T) | Logistic model (S); Decision tree (S); Random forest (S) |
| Malykh and Rudetskiy (2018) | Unclear | Unclear | Unclear |
| Zhang et al. (2018) | Gaussian process model | Complete | Weighted support vector machine (S);<br>Weighted Gaussian process regression (S) |
| N. Wang et al. (2019) | Weighted sum | Combinatorial (T) | Logistic model (S); Random forest (S);<br>Nearest neighbors (S) |
| Ma et al. (2020) | Weighted sum | Combinatorial | Extreme learning machine (S) |
| Y. Wang et al. (2020) | Weighted sum | Statistical | Linear model |
| Tang et al. (2021);<br>Ng et al. (2021) | Locally supervised metric learning (S) | Causal inference matching | Statistical test (S) |
| Doborjeh et al. (2022) | Euclidean distance;<br>Signal-to-noise ratio (S) | Statistical (T) | Spiking neural network (S) |
| Liu et al. (2022) | Weighted Manhattan distance (R,T) | Proportional (T) | Logistic model (S) |

Another interesting pattern lay in the years of publication (Figure 2). While these studies trace back to the late 1990s, they have become more common, which suggests that this is an active, though not rapidly expanding, approach. We note that CBR in health and medicine originated as early as 1990, and localized modeling emerged soon after CBR had established itself; the idea has been “in the air” for as long as CBR has been in use.

**Figure 2:**
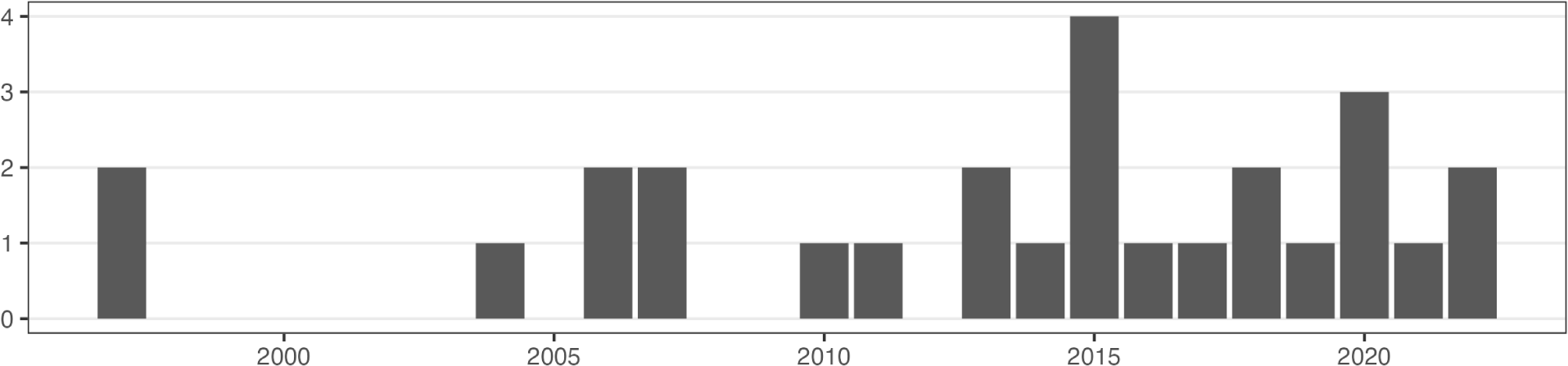
Number of publications in our sample each year.

Another dominant characteristic of included studies is their broader remit. We categorized studies as “knowledge” or “practice” based on whether their aim was to produce generalizable knowledge or to improve practice. We classified 8 of the studies as “knowledge”, making the majority of studies from a “practice” standpoint. These studies aimed to provide tools for use in clinic or to improve outcomes.

Lastly, we observed a commonality among 6 of the included studies in having evaluated methods using leave-one-out cross-validation (LOOCV). The purpose of this measure is to estimate the overall performance of certain factors when used to make predictions, particularly utilized on smaller data sets, where models benefit greatly from larger training sets and additional model fitting is less costly.

In addition to the aim of its analysis, we coded several aspects of the design of each study, including the source and type of data and the clinical task the model performed (Table 1), as well as the specific methodology and terminology adopted (Table 6). In the following subsections, we take a closer look at these design elements and their reported justifications and limitations.

**Table 6:**
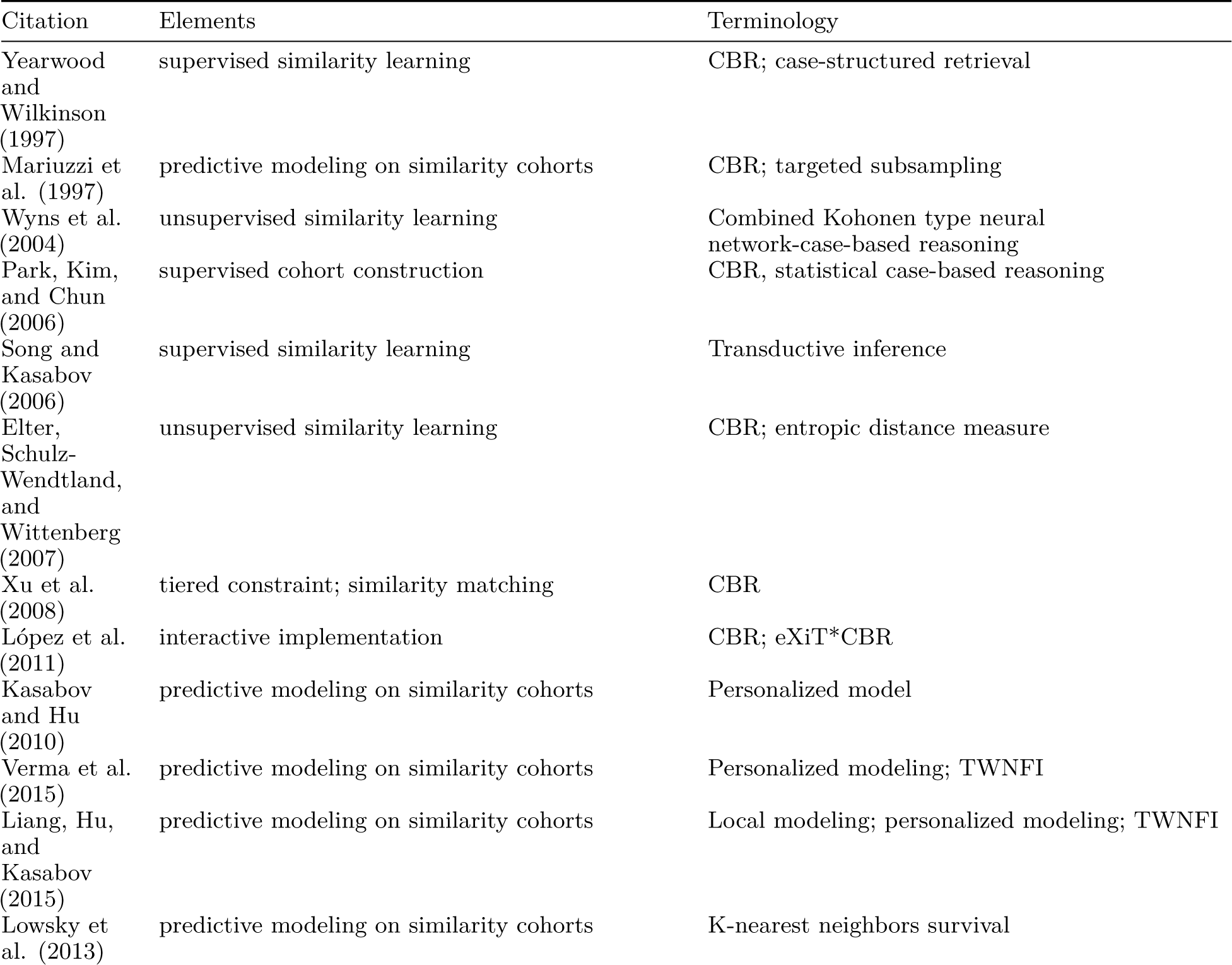
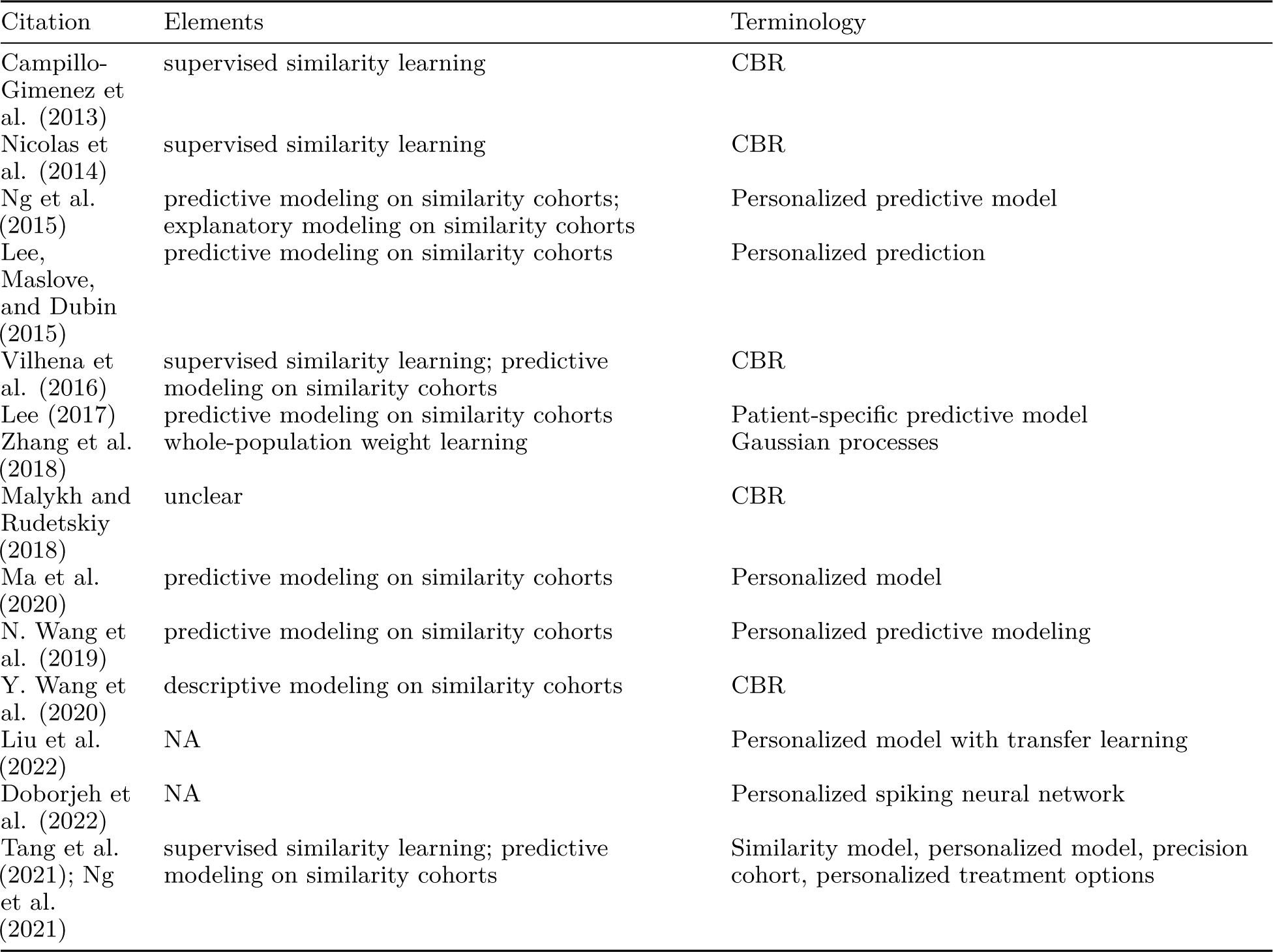
Methodological elements of studies included in the synthesis.

| Citation | Elements | Terminology |
| --- | --- | --- |
| Yearwood and Wilkinson (1997) | supervised similarity learning | CBR; case-structured retrieval |
| Mariuzzi et al. (1997) | predictive modeling on similarity cohorts | CBR; targeted subsampling |
| Wyns et al. (2004) | unsupervised similarity learning | Combined Kohonen type neural network-case-based reasoning |
| Park, Kim, and Chun (2006) | supervised cohort construction | CBR, statistical case-based reasoning |
| Song and Kasabov (2006) | supervised similarity learning | Transductive inference |
| Elter, Schulz-Wendtland, and Wittenberg (2007) | unsupervised similarity learning | CBR; entropic distance measure |
| Xu et al. (2008) | tiered constraint; similarity matching | CBR |
| López et al. (2011) | interactive implementation | CBR; eXiT*CBR |
| Kasabov and Hu (2010) | predictive modeling on similarity cohorts | Personalized model |
| Verma et al. (2015) | predictive modeling on similarity cohorts | Personalized modeling; TWNFI |
| Liang, Hu, and Kasabov (2015) | predictive modeling on similarity cohorts | Local modeling; personalized modeling; TWNFI |
| Lowsky et al. (2013) | predictive modeling on similarity cohorts | K-nearest neighbors survival |
| Campillo-Gimenez et al. (2013) | supervised similarity learning | CBR |
| Nicolas et al. (2014) | supervised similarity learning | CBR |
| Ng et al. (2015) | predictive modeling on similarity cohorts;<br>explanatory modeling on similarity cohorts | Personalized predictive model |
| Lee, Maslove, and Dubin (2015) | predictive modeling on similarity cohorts | Personalized prediction |
| Vilhena et al. (2016) | supervised similarity learning; predictive modeling on similarity cohorts | CBR |
| Lee (2017) | predictive modeling on similarity cohorts | Patient-specific predictive model |
| Zhang et al. (2018) | whole-population weight learning | Gaussian processes |
| Malykh and Rudetskiy (2018) | unclear | CBR |
| Ma et al. (2020) | predictive modeling on similarity cohorts | Personalized model |
| N. Wang et al. (2019) | predictive modeling on similarity cohorts | Personalized predictive modeling |
| Y. Wang et al. (2020) | descriptive modeling on similarity cohorts | CBR |
| Liu et al. (2022) | NA | Personalized model with transfer learning |
| Doborjeh et al. (2022) | NA | Personalized spiking neural network |
| Tang et al. (2021); Ng et al. (2021) | supervised similarity learning; predictive modeling on similarity cohorts | Similarity model, personalized model, precision cohort, personalized treatment options |

### 3.3 Application domains

While all included studies were reported in scientific and medical journals, the vast majority were oriented toward clinical practice rather than medical research. For example, a 2006 study specifically evaluated the usefulness of CBR-based explanations for the purpose of decision support (Doyle, Cunningham, and Walsh 2006). This study was part of a much larger literature on CBR systems and was included here despite relying on NN prediction because it used an unconventional voting scheme to generate recommendations. A more recent study took essentially the same focus with respect to a proposed clinical risk prediction model, which amounted to CBR with a novel weighting scheme on predictors informed by expert consensus (Fang et al. 2021). In both cases a prototype implementation was deployed in an experimental setting for evaluation.

The most common clinical motivations were individualized detection or diagnosis, prognosis or outcome prediction, and treatment or care recommendation. The plurality focused on prognosis or outcome prediction, often using time-to-event analysis: Mariuzzi et al. (1997) used CBR to predict survival time from several geometric properties of breast tumors. Lowsky et al. (2013) used CBR with non-parametric survival models on registry data to predict patient–graft survival times following kidney transplantation. Lee, Maslove, and Dubin (2015) and Lee (2017) used localized logistic regression and random forest modeling to predict 30-day mortality following discharge for ICU patients. Vilhena et al. (2016) developed a CBR cycle around a clustering-informed similarity matching procedure and an artificial neural network–based classifier to identify thrombophilia patients at high risk of thrombotic episodes. Ma et al. (2020) took a similarity cohort–based approach to predicting length of stay for ICU patients. Doborjeh et al. (2022) localized spiking neural networks in order to assess stroke risk from up to 7-day time series of clinical and environmental factors. Liu et al. (2022) incorporated global-to-local transfer learning into localized models of acute kidney injury risk in hospitalized patients. Also of note, from a public health perspective, Xu et al. (2008) used CBR to predict rehabilitation time as well as disability risk for unemployed workers experiencing chronic pain.

Toward detection and diagnosis, Wyns et al. (2004) proposed a hybrid neural net–CBR system to distinguish (with confidence bounds) arthritic versus control patients, based on several histological features. Nicolas et al. (2014) used collaborative multilabel CBR to subtype melanoma patients based on confocal and dermoscopy images. N. Wang et al. (2019) used localized models built from a multi-type additive similarity measure to distinguish type 2 diabetic versus control populations. Along the way, Song and Kasabov (2006), Kasabov and Hu (2010), and Verma et al. (2015) took an iterative model-building approach to several tasks: predicting glomerular filtration rate, a key indicator of renal function, from demographic and physiological variables; identifying patients with colon cancer using a large number of gene expression measurements; and identifying patients with type 2 diabetes based on demographic, physiological, and genetic variables.

While several studies emphasized the potential or actual value to decision-making of their methods and tools, only one incorporated treatment decisions into their approach: By taking “decision points” as their units of analysis, Tang et al. (2021) and Ng et al. (2021) built not classifiers or predictors but comparative effectiveness models into a localized framework, providing for the first time in our sample explicitly prescriptive rather than descriptive clinical decision support.

A partial exception to this focus was a 2014 study that also reported a decision support tool, in this case for early diagnosis of melanoma from clinical data and dermoscopy images (Nicolas et al. 2014). While the stated objectives were analogous, specific emphasis was placed on the acquisition of new knowledge through the development of the tool, including the systematic generation of new data and creation of a clinical ontology. This study was included for its use of a collaborative classifier that drew from multiple modeling approaches. In keeping with this focus of the included literature, the stated objectives of the proposed methods were more often (or additionally) to predict outcomes or to recommend interventions than only to diagnose disease.

The mosaic plot in Figure 3 summarizes the relation between data source, clinical task, and aim.

**Figure 3:**
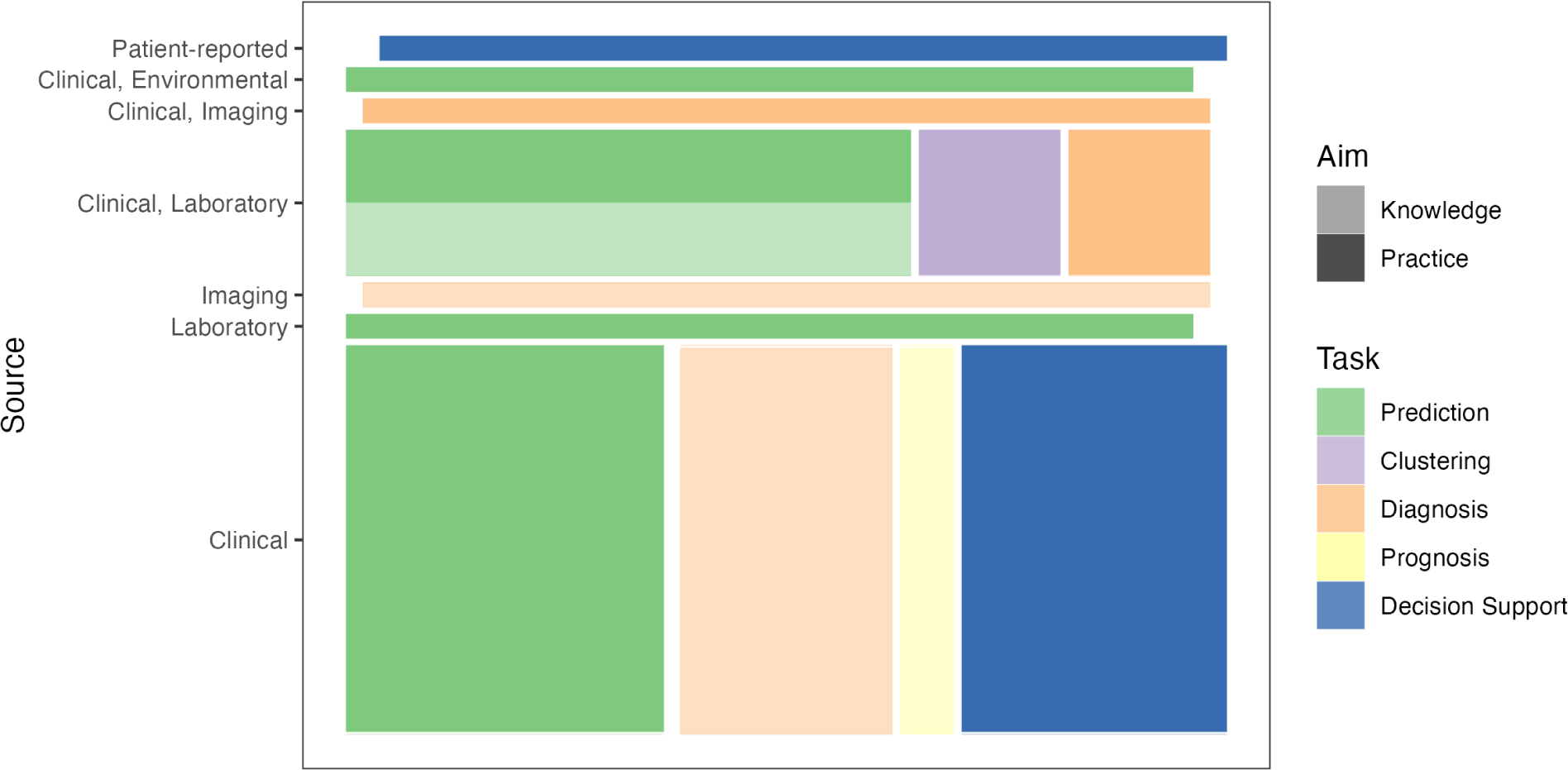
Share of studies characterized by several design elements: data source (row), clinical task (fill), and remit (opacity).

### 3.4 Rationales

The included studies hypothesized, asserted, or assumed several benefits of localized modeling specific to clinical and medical settings and advantages over other modeling approaches. The most common was that the restriction to similar or relevant past cases would improve predictive performance for the index case (Mariuzzi et al. 1997; Liang, Hu, and Kasabov 2015; Ng et al. 2015; Lee 2017). In particular, Lee, Maslove, and Dubin (2015) hypothesized and confirmed that the value of each past case would be positively related to its similarity to the index case, an assumption built in to the weighting schemes of other approaches. Lowsky et al. (2013) made a different case, that the fewer parametric assumptions and complications of a CBR-style model would allow for greater accuracy. Additionally, several investigators asserted that the use of localized cohorts befit the clinical focus on the individual patient rather than the population, without reference to performance (Song and Kasabov 2006; Xu et al. 2008).

In an interesting contrast, Tang et al. (2021) and Ng et al. (2021) argued that their localized approach using the larger and more heterogeneous populations covered by EHR-derived data could better capture the messy and diverse lessons of everyday practice, as a counterpart to guidelines based on randomized controlled trials. Their emphasis on the enabling role of EHRs to power methods well-suited to large, structured data repositories was shared by several others (Campillo-Gimenez et al. 2013; Nicolas et al. 2014; Verma et al. 2015; Lee 2017). Verma et al. (2015) and Zhang et al. (2018) additionally pointed out that localized models are adaptable to noise in the data, variation in patterns of missingness, and (harkening to the last step of the R4 cycle) addition of new cases with known outcomes to the corpus. These properties, they said, tend to be more difficult for whole-population models to handle.

The other frequent advantage attributed to localized modeling was interpretability. Elter, Schulz-Wendtland, and Wittenberg (2007) and Nicolas et al. (2014) emphasized the value of the detailed past cases, available to the user, on which predictions are based. This “self-explanation capability” made outputs more intelligible to physicians in the CDS setting. Ng et al. (2015) additionally pointed out that localized GLMs yield localized effect estimates, which may help investigators identify individually relevant risk factors. Liu et al. (2022) took this idea further, systematically comparing regression coefficients for specific risk factors across dozens of diagnostic subpopulations. Doborjeh et al. (2022) used localized time series models to identify environmental changes associated with increased stroke risk.

The remaining coded rationales were for augmentations or hybridizations of then-conventional CBR, most of which defended the use of other tools to improve performance via cohort selection (Campillo-Gimenez et al. 2013; Nicolas et al. 2014; Vilhena et al. 2016) or to make models and outputs more interpretable (López et al. 2011; N. Wang et al. 2019). Liang, Hu, and Kasabov (2015) argued for simultaneous optimization of cohort and feature selection with model parameterization; their TWNFI approach combines localization with regularization.

### 3.5 Challenges

CBR can be understood as the opposite side of a trade-off with rule-based reasoning between model size and model complexity: Doyle, Cunningham, and Walsh (2006) describe their approach as “knowledge-light”, in that “the cases do not contain explicit explanation structures; instead, explanation is achieved by comparison of the query case with retrieved cases”. This means that the greatest performance and efficiency challenges in CBR have to do with the retrieval and revision phases in the R4 cycle. Several studies addressed these challenges: Park, Kim, and Chun (2006), while excluded from the synthesis, were the earliest to propose that cohorts be bounded by a similarity threshold rather than by the number of cases, which improved performance in their experiments. Campillo-Gimenez et al. (2013) leveraged predictor weights obtained from logistic regression to inform the similarity calculation used in retrieval. Lowsky et al. (2013) used non-parametric models to reduce the computational burden of revision (prediction). Ma et al. (2020) proposed to improve efficiency along the entire R4 cycle, but in particular by only executing task-dependent steps in real time (“just-in-time learning”, JITL), and Ng et al. (2021) and Tang et al. (2021) partitioned multiple phases in the R4 cycle into offline and online components, only the latter of which would be performed in real time as new data are received. Liu et al. (2022) used transfer learning from globally-fitted models to improve the efficiency of localized models.

Other studies addressed limitations of available tools. López et al. (2011) implemented a comprehensive CBR tool in response to the lack of general-purpose software, to enable coupling with other tools as well as to expedite development, experimentation, and uptake. Several other teams also set out to develop more generalizable and data-agnostic clinical support tools (Elter, Schulz-Wendtland, and Wittenberg 2007; Liang, Hu, and Kasabov 2015; Ng et al. 2015; Zhang et al. 2018). However, more informatical and legal challenges to implementation, including interoperability of systems and data and regulations concerning security and privacy, were rarely addressed. We discuss this further in Section 4.5.3.

Figure 5 compares the rates at which several methodological elements are invoked in the sample.

### 3.6 Evaluations

Most included studies quantitatively compared the predictive performance of their proposed method(s) to one or more comparators. What we took to be the signature results are collated in Table 7 in the Appendix. Note that we exercised some judgment in classifying methods as proposals and comparators, as in some cases all methods were original but only some showcased main ideas. It would be impractical to meta-analyze these numbers due to the great variety of settings, problems, data types, techniques, and choices involved.

**Table 7:**
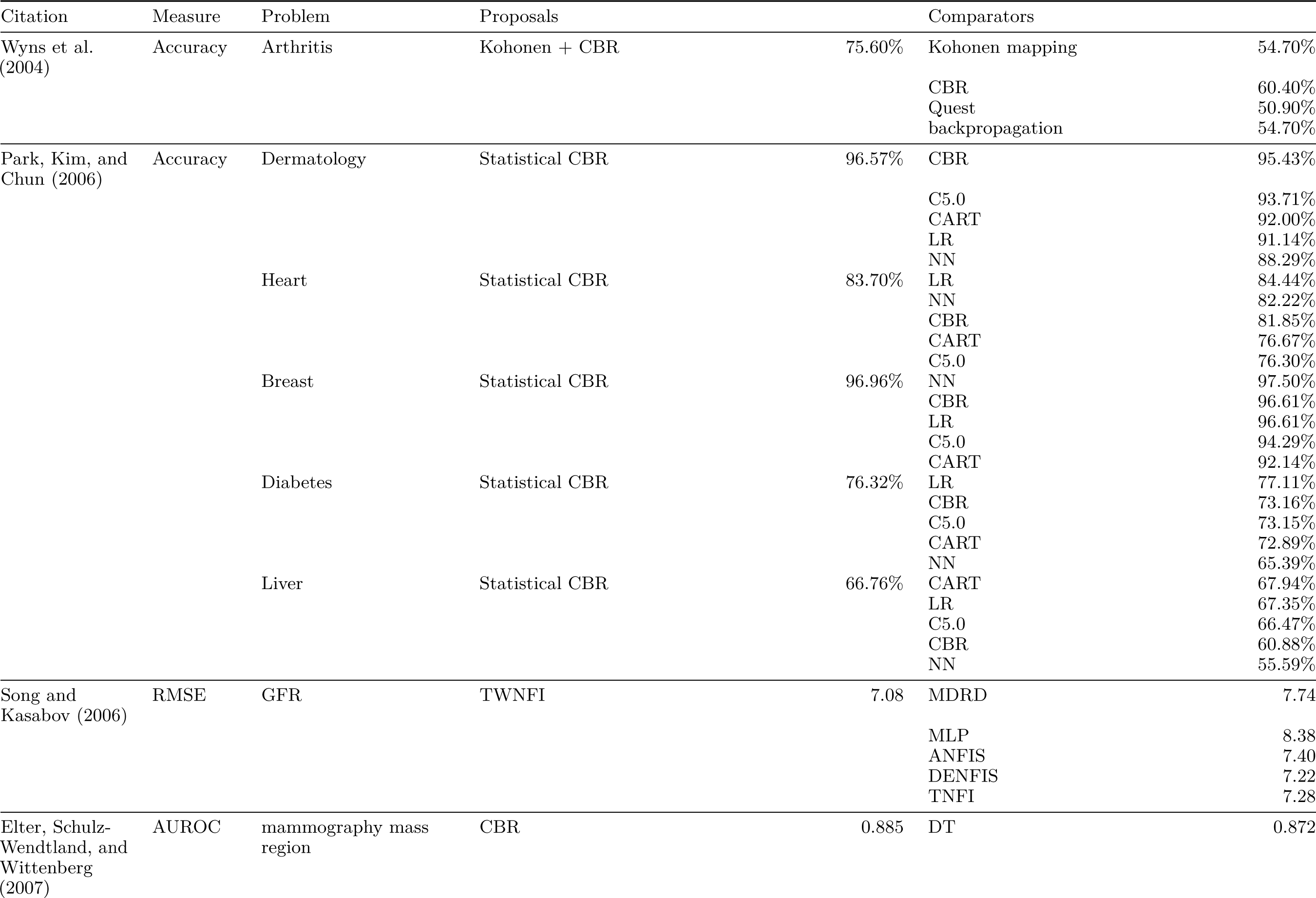
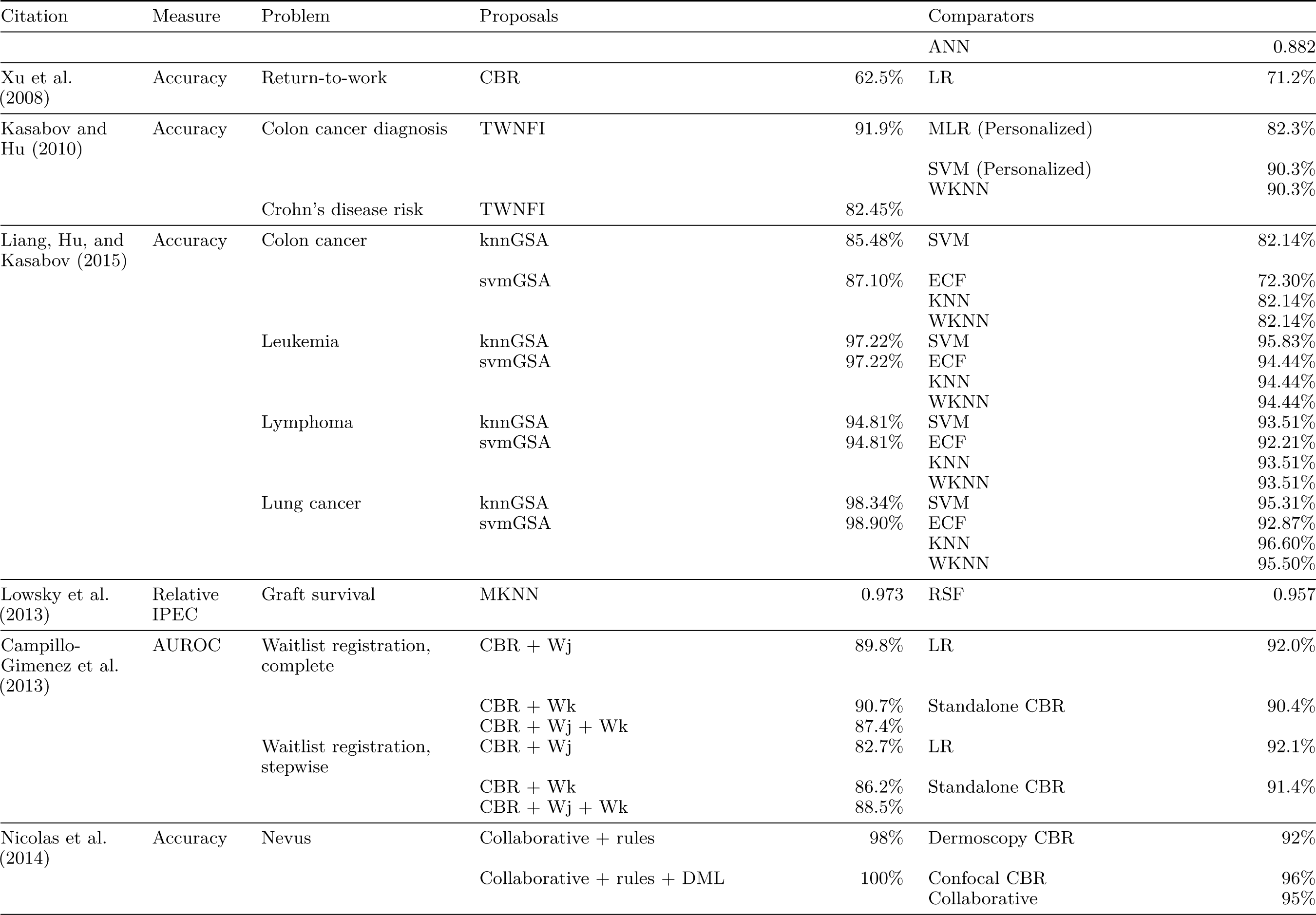
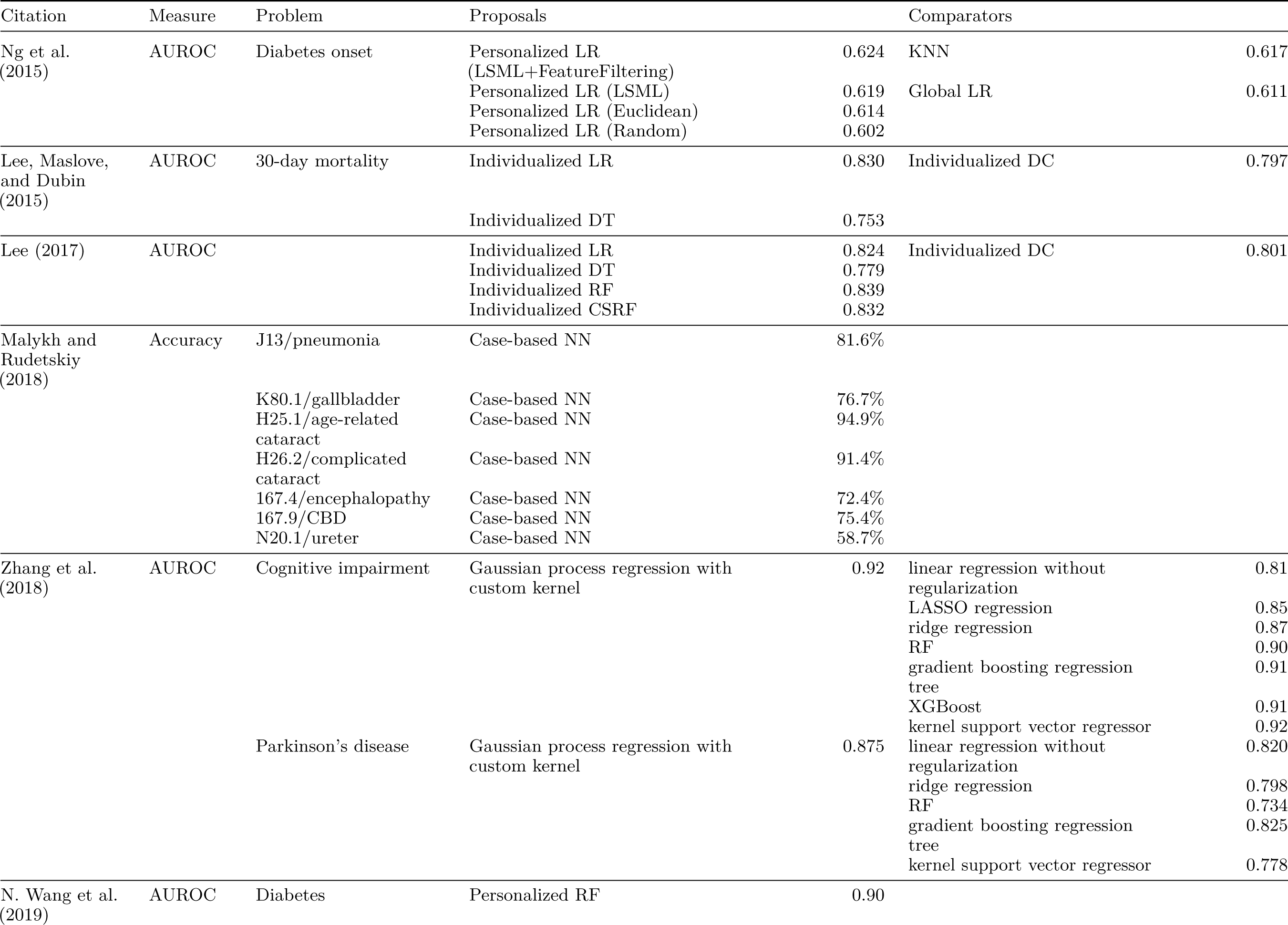
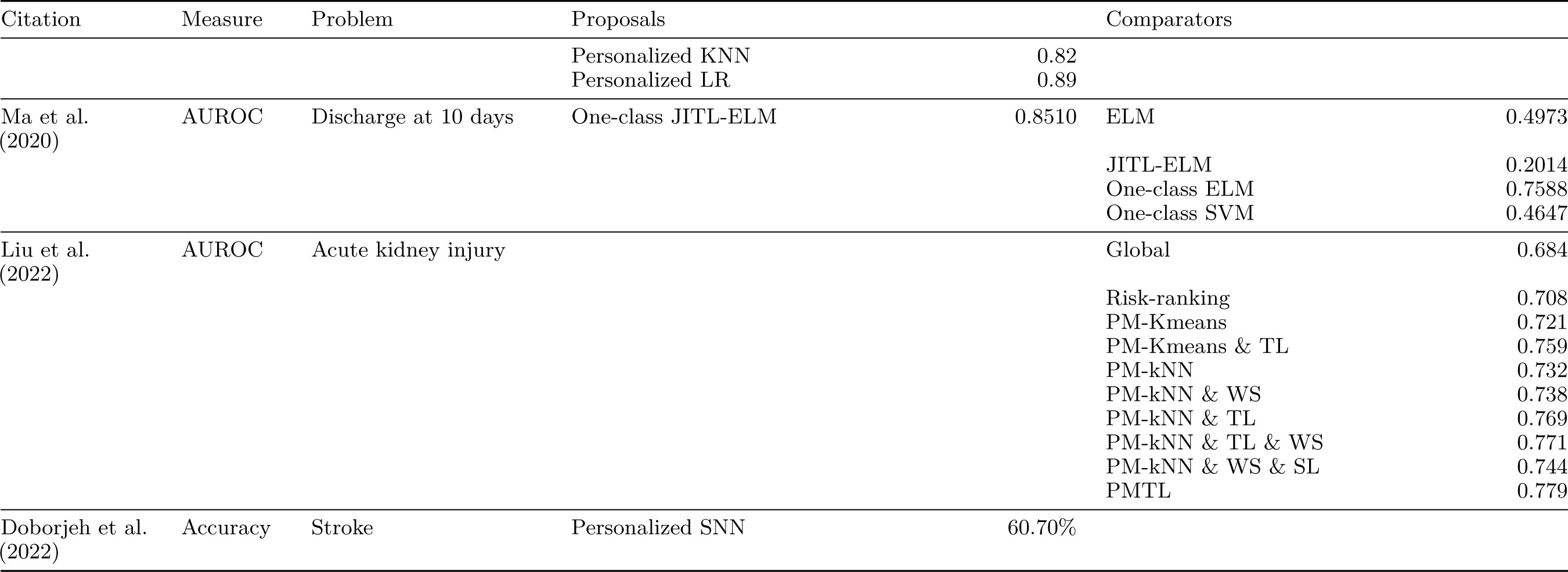
Evaluations of proposed methods and comparisons to alternative methods. (See original studies for full names and descriptions.)

We do observe one clear pattern: All of the proposed methods that most evidently outperformed their comparators—Kohonen + CBR (Wyns et al. 2004), TWNFI Kasabov and Hu (2010), gravitational search algorithms (GSA) (Liang, Hu, and Kasabov 2015), CBR + rules (+ DML) (Nicolas et al. 2014), Gaussian process regression (Zhang et al. 2018), JITL-ELM (Ma et al. 2020)—are hybrids of localized modeling (in some cases CBR) with other techniques, often DML. Though Campillo-Gimenez et al. (2013) and Ng et al. (2015) report non-superior performance by such hybrids, the pattern suggests the importance of the similarity measure to the retrieval step. Meanwhile, when proposed approaches targeted cohort demarcation or choice of predictive model—statistical CBR (Park, Kim, and Chun 2006); individualized logistic regression, decision tree, and random forest (Lee, Maslove, and Dubin 2015; Lee 2017)—they did not consistently outperform comparators.

### 3.7 Identified needs

The study authors focused their recommendations and their own plans for future work mostly on technical improvements and evaluations (Mariuzzi et al. 1997; Yearwood and Wilkinson 1997). Urged improvements included full or partial automation of predictor selection (Mariuzzi et al. 1997; Yearwood and Wilkinson 1997), similarity learning (Mariuzzi et al. 1997; N. Wang et al. 2019), and parameter optimization (Song and Kasabov 2006; Lee 2017); extensions to new data structures (López et al. 2011), data types (Liang, Hu, and Kasabov 2015; Verma et al. 2015; Malykh and Rudetskiy 2018), and reasoning systems (Nicolas et al. 2014); and the use of more advanced model components to improve accuracy or efficiency (Lowsky et al. 2013; Lee, Maslove, and Dubin 2015; Liang, Hu, and Kasabov 2015; Zhang et al. 2018). Authors also urged validations and independent evaluations using larger or more comprehensive data sets (Elter, Schulz-Wendtland, and Wittenberg 2007; Xu et al. 2008; Verma et al. 2015; Ng et al. 2015), using data aggregated from multiple health systems (Lee, Maslove, and Dubin 2015; Lee 2017; Tang et al. 2021; Ng et al. 2021), and in other care settings or disease contexts (Song and Kasabov 2006; Zhang et al. 2018; Tang et al. 2021; Ng et al. 2021).

Less common were calls to strengthen the connection between the methods and the users. Some authors urged the incorporation of (human-derived) domain knowledge into the data or models (Yearwood and Wilkinson 1997; N. Wang et al. 2019). Others suggested new uses of their modeling approaches: to measure feature importance (Wyns et al. 2004), to make models more expressive (Lee, Maslove, and Dubin 2015), and to combine information obtained both from global and from localized models (Ng et al. 2015). Most of this incremental work was indeed carried out in later investigations in our sample. That said, we did not find reports of successful implementations of these tools in clinical practice. (Several excluded studies did showcase uses of CBR in practice.)

## 4 Discussion

After commenting briefly on our review process, we draw several observations from our encoding of the included studies. We first focus on their methodological choices and innovations, then on their conventions and language. We spend most of the section discussing the differences and commonalities of the techniques used, toward a general method that encompasses most or all cases. We then suggest some avenues for future studies and conclude with our key takeaways.

### 4.1 Process

The low inter-rater reliability was due, in parts, to different interpretations of some eligibility criteria by the authors, inconsistent terminology across the sample, and incomplete reporting of resources and methods in the sample. Regarding interpretation of criteria, some wording of the criteria was adjusted following discussions between the reviewing authors during full-text review to better specify an agreed-upon meaning. We will discuss the different domains, terminologies, and reporting issues of the sample in the remainder of this section.

### 4.2 Study designs

Consistent with their orientation, almost all included studies were conducted using clinical data, only occasionally together with patient-reported (2), laboratory (3), image (1), and environmental (1) data. We suggest three reasons for this: First, this literature traces back to the 1990s, before -omic data could be generated cost-effectively at scale. Second, CBR in particular has a strong tradition in clinical decision support, where the focus of our sample remains throughout the review period. Third, because most -omic data are highly homogeneous—all measurements are made along or can be transformed to a common scale, e.g. greyscale pixellations for X-ray images and transcripts per million for RNA-seq data—more deeply theoretical analysis techniques have been developed and come into wide use. While variations on correlation-based approaches like EHR-based phenome-wide association studies and similarity-based methods like CBR itself have been developed, more mechanistic and probabilistic tools have not become a domain standard.

Because we excluded studies that used conventional NN prediction, many included studies reported new approaches to adaptation subsequent to retrieval. Very few proposed novel similarity measures, possibly because larger studies tended to be reported in separate articles detailing experiments with specific components of the process. However, this also suggests that few experimental studies have focused on the unified development of new retrieval and adaptation strategies. We also note that the majority of studies evaluated and compared methods using leave-one-out cross-validation (LOOCV). This is an appropriate technique when available data are scarce, and indeed most data sets used by included studies numbered in the hundreds of cases or fewer. This likely follows from the older age of many included studies and from their consistent primary focus on clinical data, which is more costly to collect and comes with more restrictions on its use.

### 4.3 Coherence

The motivational and methodological unity of these studies does not reflect a unified research program. Besides the lack of any primary journal of record, we observed collaborations only among the authors of smaller contiguous programs, including applications of the TWNFI methodology (Song and Kasabov 2006; Verma et al. 2015), individualized mortality prediction for ICU patients (Lee, Maslove, and Dubin 2015; Lee 2017) and the use of more explanatory models to prioritize predictors or treatments for chronic disease (Ng et al. 2015; Tang et al. 2021; Ng et al. 2021).

These programs used varying terminology for common concepts, and no common term is in use for what we here refer to as localized modeling; authors described their approaches as “targeted prognosis” (Mariuzzi et al. 1997), “transductive inference” (Song and Kasabov 2006), “personalized decision support” (Lee, Maslove, and Dubin 2015), “personalized (predictive) models” (Liang, Hu, and Kasabov 2015, in contrast to “local models”; Ng et al. 2015; N. Wang et al. 2019; Ma et al. 2020; Liu et al. 2022; Doborjeh et al. 2022), and “precision cohort” analysis (N. Wang et al. 2019; Tang et al. 2021; Ng et al. 2021). We find uses of “targeted”, “personalized”, and “precision” generic and imprecise, while transductive inference is an established term for a broader set of methods in ML. Because *local* naturally contrasts with *global*, we propose “localized models” as a suitable term for this counterpart to globally-fitted models.^2^

### 4.4 Synthesis

The 27 studies included in our synthesis used composite techniques that we organized into two major types. One type, *similarity learning*, was used in 8 studies that tie in to the much larger literature on DML (Yang 2006). The other, *cohort thresholding*, focused on choosing or optimizing the manner in which cohorts were demarcated using the similarity measure.

#### 4.4.1 Similarity learning and threshold optimization

The similarity learning approaches took a variety of forms. These techniques are designed to detect structure in data, especially associations between predictors and responses, and to use this structure to inform the definition of a patient similarity measure. Eight studies used response values in training data sets to supervise similarity learning while two used unsupervised learning (Table 6). Based on the key properties of DML algorithms identified by Bellet, Habrard, and Sebban (2014), most learned measures were non-linear while some were locally learned, and some were optimized globally while others locally. Not all qualified as DML, since the resulting measures would not necessarily satisfy the triangle inequality. Each used its similarity measure to retrieve training cases relevant to each testing case. Once retrieved, most took a standard NN approach to generating predictions; two exceptions (Vilhena et al. 2016; Tang et al. 2021) fit predictive models to the retrieved cohorts.

Several studies used unsupervised similarity learning, for example the Mahalanobis distance (Lowsky et al. 2013) and the Kolmogorov entropy-based distance (Elter, Schulz-Wendtland, and Wittenberg 2007). In particular, Yearwood and Wilkinson (1997) defined similarity as a weighted sum of differences in predictor values and used linear regression to optimize the weights for the predictive accuracy of the cohort they retrieve. Others used supervised learning: Song and Kasabov (2006) proposed an iterative algorithm to optimize a set of fuzzy inference rules used to retrieve relevant cases and generate a prediction, using back-propagation on the rules’ parameters. Their method was used in several later studies returned by our search, which are not discussed here because they did not originate the technique. Nicolas et al. (2014) explicitly appeal to a supervised distance metric learning (DML) technique (Xing et al. 2002) to obtain a similarity measure on their set of dermoscopy and confocal images that most effectively separates malignant from benign melanoma tumors. We will discuss the remaining uses of supervised similarity learning shortly.

Distinct from but related to similarity learning was supervised cohort construction. This was an adaptive step taken by Park, Kim, and Chun (2006) to optimize the predictive performance of models fitted to cohorts retrieved using a fixed similarity rather than count threshold, a counterpart to conventional CBR they called “statistical CBR”.^3^ The step used a heuristic procedure to locate a similarity threshold that (locally) maximizes predictive accuracy on the training set. This is analogous to optimizing the neighborhood size parameter in NN predictive modeling, so we will not discuss it further except to mention that the authors found statistical CBR to outperform conventional CBR on several data sets. Campillo-Gimenez et al. (2013) combined these learning techniques: They used logistic regression on the training set to obtain weights for the predictors (by predicting the binary outcome of kidney transplant waitlist registration) and for the cases (by predicting agreement of outcome between an index case and other training cases), and they used exhaustion to optimize the size of the retrieved cohort for the accuracy of the NN model using the previously optimized weights.

As noted above, these instances of similarity learning fit into a much larger literature on DML, which has seen widespread use in health informatics and modeling. That these studies satisfied our review criteria is due to the novelty of the techniques at the time of publication and the detailed attention paid to their techniques by the authors. It also reflects a limitation of our study and of the literature we set out to retrieve: We are aware of no standard terms in use to identify the family of techniques that fit statistical models to similarity-based cohorts. The retrieved studies whose use of such techniques alone satisfied our criteria variably termed them individualized, personalized, local, and patient-specific models, among other terms (Table 6). We prefer the term localized models, which is both sensitive and specific to uses of a similarity or distance measure to retrieve a cohort to which a model is fit: It includes some properly included techniques we did not have in mind (Vilhena et al. 2016) yet excludes other techniques commonly referred to as individualized, personalized, or precision.

#### 4.4.2 Localized models

In addition to Lowsky et al. (2013), Liang, Hu, and Kasabov (2015), Ng et al. (2015), Lee (2017), Vilhena et al. (2016), Tang et al. (2021), and Ng et al. (2021), seven other studies employed localized models: Mariuzzi et al. (1997), Lee, Maslove, and Dubin (2015), Verma et al. (2015), N. Wang et al. (2019), Ma et al. (2020), Liu et al. (2022), and Doborjeh et al. (2022). In most cases these models were purely predictive in application; of the exceptions, Mariuzzi et al. (1997) produced purely descriptive models of survival outcomes, which were evaluated for their precision rather than their accuracy, while Ng et al. (2015) used generalized regression models descriptively as well as predictively, as did Tang et al. (2021), Ng et al. (2021), and Liu et al. (2022) later. The most common approach to cohort selection was to optimize a size threshold via manual exploration (Mariuzzi et al. 1997; Lee, Maslove, and Dubin 2015; Ng et al. 2015; Lee 2017; N. Wang et al. 2019; Vilhena et al. 2016) or via cross-validation (Lowsky et al. 2013; Verma et al. 2015). Further in the former direction, one study (Ma et al. 2020) set a fixed cohort size while two (Liang, Hu, and Kasabov 2015; Tang et al. 2021; Ng et al. 2021) devised more sophisticated algorithms to balance multiple desiderata including predictive accuracy.

Several themes emerged from this sample. Foremost was the optimization of retrieved cohort sizes for some measure of model performance. Despite the early recommendation by Park, Kim, and Chun (2006) to threshold cohorts by similarity rather than by cardinality, only the one previously published study (Mariuzzi et al. 1997) and the most recent study (Tang et al. 2021; Ng et al. 2021) in this corpus took this approach, though Liu et al. (2022) took a third option by retrieving fixed proportions of training data. Viewed as a hyperparameter of the individualized modeling approach, cohort size can be treated in the same way as neighborhood size in NN prediction (viewed here as a special case of localized modeling wherein the model is a simple summary statistic), so we will not discuss it further.

An alternative approach was to optimize multiple hyperparameters together: Kasabov and Hu (2010) proposed an iterative algorithm to settle on an optimal number and set of predictors as well as number of training cases (comprising the retrieved cohort) that was later used by Liang, Hu, and Kasabov (2015). More recently, Tang et al. (2021) and Ng et al. (2021) proposed a three-step process for cohort selection that filtered by exact match for one subset of predictors, used domain-informed similarity measures to rank these, and optimized the similarity threshold for a trade-off between cohort size and a measure of bias called cohort balance. Notably, though their similarity measure was supervised, this optimization process was not.

With the exception of the descriptive survival models of Mariuzzi et al. (1997), only one thread in this corpus concerned itself with the interpretability of localized models: Ng et al. (2015) fit logistic regression models to similarity-based cohorts and examined not only their predictive performance but the localized sets of largest and most detectable predictors, termed risk profiles, and how they differed across the testing population. This approach informed their later use of localized comparative effectiveness–style models to recommend courses of treatment at decision points during a monitored patent’s stay (Tang et al. 2021; Ng et al. 2021), and it was later used by Liu et al. (2022) to show a dependency of predictor importance on the patient disease subgroup. These studies suggest much wider potential for localized real-world evidence generation, which has long been promoted as a promise of advances in clinical and health informatics (Longhurst, Harrington, and Shah 2014).

#### 4.4.3 A general framework

This relatively small sample exhibited a wide range of approaches, both technically and conceptually. Discussion of the technical diversity of these approaches has been summarized above and published in greater detail in previous reviews (Choudhury and Begum 2016; Sharafoddini, Dubin, and Lee 2017; Parimbelli et al. 2018), and we focus here on the conceptual. The design of a localized modeling approach can be decomposed into three largely independent choices: (1) how to measure the similarity between cases, (2) how to retrieve a cohort of cases similar to an index case, and (3) how to generate a prediction or other statistical insight for the index case from the similarity cohort. The choices are the *similarity measure*, the *cohort retrieval*, and the *statistical model*, respectively. These are diagrammed in Figure 4.

**Figure 4:**
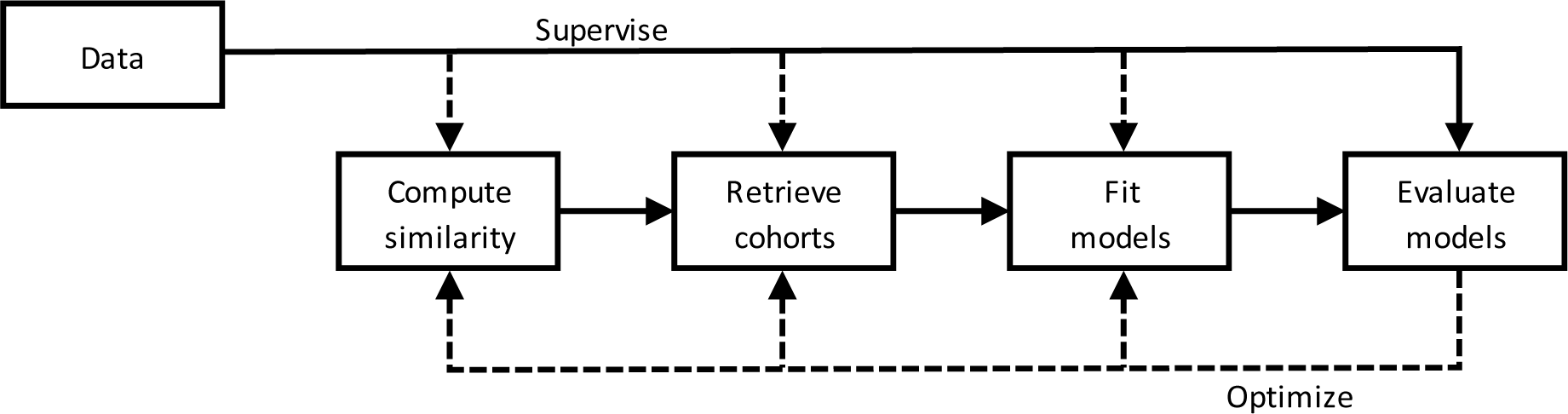
General framework for localized models. Dashed arrows indicate optional steps.

Each step can be unsupervised supervised: Unsupervised similarity measures include discrete measures like the Levenshtein distance and continuous measures like cosine similarity, while supervised measures include the Mahalanobis distance and random forest proximity as well as several composite measures with weights calculated from the data. Most retrieval steps obtained cohorts with a uniform size (cardinality) or similarity threshold, and the remaining were likewise only informed by predictors; thus, while a retrieval step could in principle be supervised, in this sample none were. Almost all models were supervised, being that most performed predictive tasks, though Mariuzzi et al. (1997) modeled only survival rates within localized cohorts. Meanwhile, each step can be optimized in a ML fashion by having its parameters tuned to improve performance. We found one study that tuned the calculation of similarity (Liu et al. 2022), though most uniform cohort sizes were also tuned. Some earlier studies tuned generalized regression models, but more recent studies did not.

### 4.5 Directions and expectations for future work

#### 4.5.1 Performance

As discussed in Section 3.4, most studies were premised on the potential predictive value of localized models. As cautioned in Section 3.6, not all experiments affirmed this premise. Most of those that did involved using metric learning to improve retrieval and sometimes iterative optimization of the metric and of the localized model. Some studies showcased results using a variety of such specializations (Campillo-Gimenez et al. 2013; Ng et al. 2015; Zhang et al. 2018; Liu et al. 2022). However, because these advanced implementations were mostly compared against their precursors or commonplace alternatives, we cannot speak to their performance or any trade-offs with respect to each other. This would be a worthy goal of future work.

The value of this work would be enhanced by standardization and modularization. First, studies using localized models should locate the proposed approach and any variations in a shared specification space. For this purpose, we propose the framework outlined above, at least as a starting point. When a specification invokes complex techniques, for example supervised DML or neural network classifiers, it should be compared at least to alternative specifications using simpler alternatives. Campillo-Gimenez et al. (2013) and Liu et al. (2022) provide excellent examples in which all possible choices are toggled and yield a neatly nested comparison. Additionally, some state-of-the-art models, including others that obtain individualized predictions, should be included in comparisons. Finally, implementations should be not only published on public repositories but designed in such a way that users familiar with the underlying language can substitute a technique of their choosing for any part of the specification (similarity, retrieval, fitting, evaluation). We believe these principles will make reported results clearer and reproduction and extension easier.

#### 4.5.2 Interpretability

While most studies identified outstanding technical needs (Section 3.7), few emphasized the need to assess human-focused qualities like user interface and user experience, interpretability, meaningfulness, or trust. Most studies identified interpretability as an advantage of localized models, though none evaluated model interpretability and few proposed new interpretative uses. Lack of trust in ML tools is a long-recognized problem that direct intepretability of model components could help alleviate, but this is more often assumed than demonstrated.

One valuable direction for future work would be to measure the utility of interpretable components in research and in clinical practice and the correctness and confidence of users in their interpretations. This is a necessary precondition for practical use but would also be a valuable contribution to the experimental literature. Another would be to compare the localized predictor importance measures obtained from localized models to the local importance measures used to explain predictions made by opaque models. This would test both measures for concurrent validity, while differences between them would inform what settings or cases are poorly served by one or the other.

#### 4.5.3 Feasibility

Much of the reviewed work was motivated by the need for modeling paradigms that perform well on a variety of tasks and in a variety of settings. This need demands not only methodologies but also architectures that are robust, versatile, and compliant in the face of diverse data models and use restrictions, yet no studies in our sample addressed the challenges of system interoperability and regulatory requirements head-on.

CBR and, by extension, localized modeling are especially susceptible to these challenges, as corpora of past cases must be aggregated from multiple institutions and from multiple systems within institutions, and models built on them must then be applicable to out-of-box data structures as well. Individual systems exhibit many dimensions of incompleteness (Weiskopf et al. 2013) and vary along other dimensions of quality (Kohane et al. 2021), and their aggregation for modeling purposes depends on several kinds of interoperability (Weber 2015). Successful deployment also depends on satisfying regulatory regimes designed to protect the privacy of patients, the security of communications, and trust between parties (Haendel et al. 2021). While most studies in our sample focused on improving care, increasingly many over time focused on generalizable knowledge. Therefore, as studies using localized modeling shift from proof of concept to feasibility, they will also need to demonstrate practical and regulatory feasibility.

#### 4.5.4 Customizability

The primary goal of these studies was to establish that localized models, and certain strategies within this paradigm, perform at or above the level of other predictive models or strategies. From a functionalist approach to reproducibility (Matarese 2022), then, later studies were broadly successful at reproducing earlier studies, and later studies provide sufficient detail to support ongoing reproduction efforts—though these might be limited by the lack of open-source code or public implementations. However, the immediate goal of most studes was to improve care, and an implied need, often enjoined but neither obtained nor reproduced, was a demonstration of practical use.

We posit that an essential component of practical usefulness is the ability of the user community to exert some control over the models. Given the impact on performance of the choice or optimization of the similarity measure, an important target for user input would be the importance of certain variables in the calculation of similarity, with an understanding of how it can impact not only the performance of the predictions but also the cohort retrieved for the model. For one example, a user may want to minimize the weight of rare diseases in medical history in order to retrieve a population with more such cases in order to better measure their associated risk to the outcome. In contrast, they may want to increase the weight of the indicating diagnosis in order to allow fewer patients from similar but distinct populations to influence the model. For another example, a user may want to down-weight socioeconomic variables like race–ethnicity in order to ensure a more diverse modeling cohort. López et al. (2011) took an important step in this direction with an adjustable and adaptable implementation. Future purely quantitative work could assess whether similarity-tuning can achieve these ends more flexibly than strict inclusion/exclusion criteria.

### 4.6 Conclusions

We propose the term *localized modeling* to encompass an approach derived from CBR in which parameterized models are fitted in a standardized way to nearest neighborhoods of past or training cases according to a measure of patient similarity. We conducted a systematic search for studies that apply localized models to tasks involving health data and synthesized these largely independently developed approaches into a general framework. While the search was limited by low inter-rater reliability and failure to recover several motivating examples, the included studies used many of the same underlying tools to build, optimize, and evaluate their methods. We therefore believe that our framework can serve to taxonomize ongoing work of this type and inform the development of customizable implementations. Indeed, the availability of increasing computational power, the diversity of tasks to which these models were applied and of technical specifications they employed, and the apparent lack of any multi-group research program to date suggest great potential for growth. Whereas precious few of the reviewed studies used these models for any task other than prediction, despite widespread suspicion among clinicians of “black-box” models and growing interest in interpretable alternatives, we recommend that future work put greater emphasis on the development and validation of interpretable localized models and on their reception by communities of medical research and clinical practice.

## Data Availability

All data generated over the course of the study is freely accessible at links provided in the manuscript.

https://www.zotero.org/groups/5017571/imsr/

https://docs.google.com/spreadsheets/d/1tpWMhYH2pyRT55K7n2J2XFs-kEV_JTuCDmXzJ4BBgNo/

https://docs.google.com/spreadsheets/d/1xvDJwiLBoI2oz8fxHJ5MjNmiju_RAlK7RJv-wXe1DAs/

https://github.com/corybrunson/imsr

## Appendix

### Research question

How is patient similarity–based individualized modeling conducted using retrospective data?

### Purpose of review

1. Provide a summary of individualized models to date.
2. Lay the groundwork for conducting a comparison study of individualized models.
3. Provide a framework for future individualized modeling studies.

### Search design

The procedure for formulating the search began with an evaluation of the research question. We highlighted specific elements within our topic of interest that we found critical to our search and listed them using an OR of ANDs, pairing terms we believed would provide our desired result. Following the solidification of the search, a thesaurus was created in which each term was expanded by synonyms that are similar enough to our core term to be applicable to our search. The expanded search string was then evaluated using the PubMed Advanced Search platform. We initially included each term and their synonyms separately to evaluate what resulted. Several terms were eliminated due to PubMed classifying them as “phrases not found” and other terms were removed to reduce the number of results, providing a more concise list of results. At the conclusions of this process for each individual term, we combined each search term using an OR of ANDs to ultimately form our search.

The search will have been designed to recover studies of the kind reviewed by the review papers from which we obtained our “seed set”. To validate the final search design, we will determine how many of the papers in this seed set that are indexed by PubMed are actually recovered by our search. In most cases, the focus of a review paper is different from ours, so we will only perform this validation test on the seed set obtained from two review papers that are (a) closest in focus to ours and (b) use terminology associated with the two distinct sub-literatures relevant to our focus: Choudhury & Begum (2016), which focuses on case-based reasoning in medicine, and Sharafoddini, Dubin, & Lee (2017), which focuses on patient similarity–based prediction models on health data. The proportion of each PubMed-indexed seed set that is recovered from our PubMed search provides a rough and optimistic yet useful estimate of the proportion of the relevant literature that our full search strategy will recover.

Once we have finalized the search as a logical pattern, we will take the following steps to generate the sample/corpus of literature that will be the starting point for our selection process.

1. The logical pattern will be converted to a search string using the syntax appropriate to each database in our search strategy. These include PubMed (already done as part of the search design), Web of Science, Academic Search Premier/Elite, and Mathematical Reviews.
2. The search will be conducted on each database and the results organized into a Zotero collection, with one subcollection for each database.
3. Duplicate results will be identified and merged. (A result obtained from multiple databases should have only one Zotero entry but should be filed under the subcollection for each database in which it was found.)

Following discussion among AC, PMJ, and JCB, we discarded results from Google Scholar due to missing abstracts, missing URLs, high overlap with other search results, and irreproducibility of the search process.

### Search strings

Here we reproduce the search strings and platform specifications used in our literature search. We first finalized the **PubMed** search string below:^4^

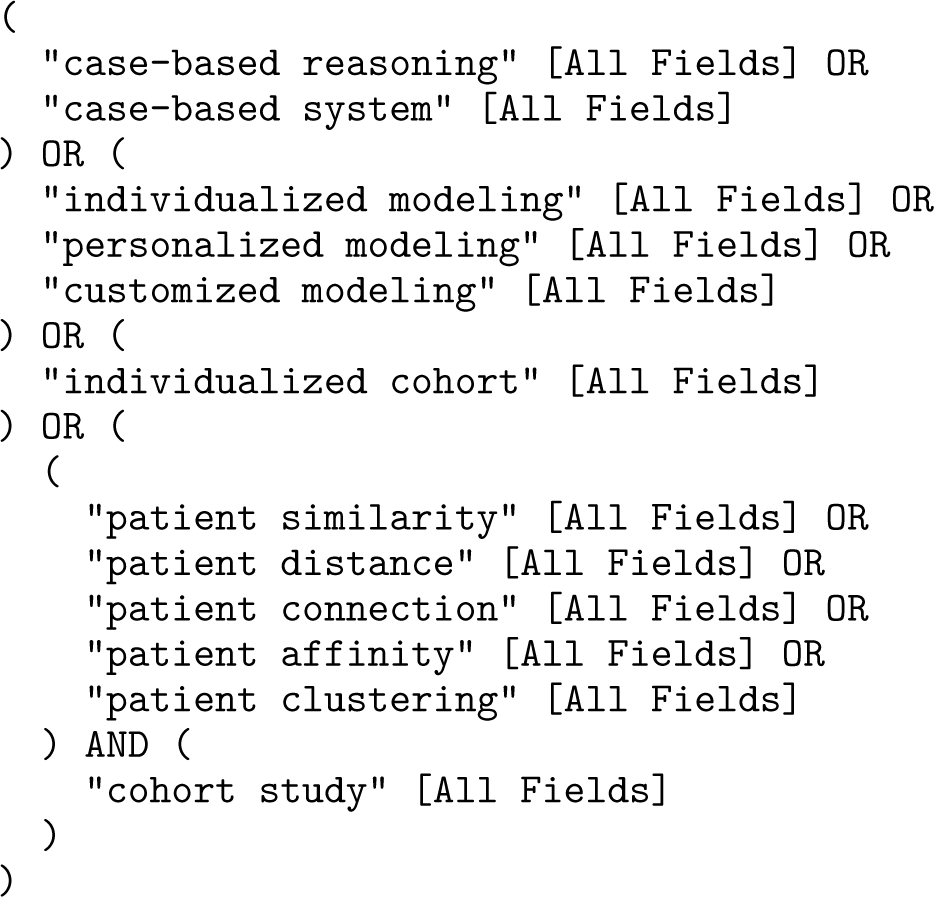

This search yielded 423 results.

We then generated analogous search strings or search strategies for the Web of Science, Academic Search Premier, and MathSciNet platforms, based on the logics and syntaxes of their respective interfaces.

For **Web of Science**, we searched for several separate disjunctions derived from the PubMed search string. The separate search strings and the number of results obtained using each are below.

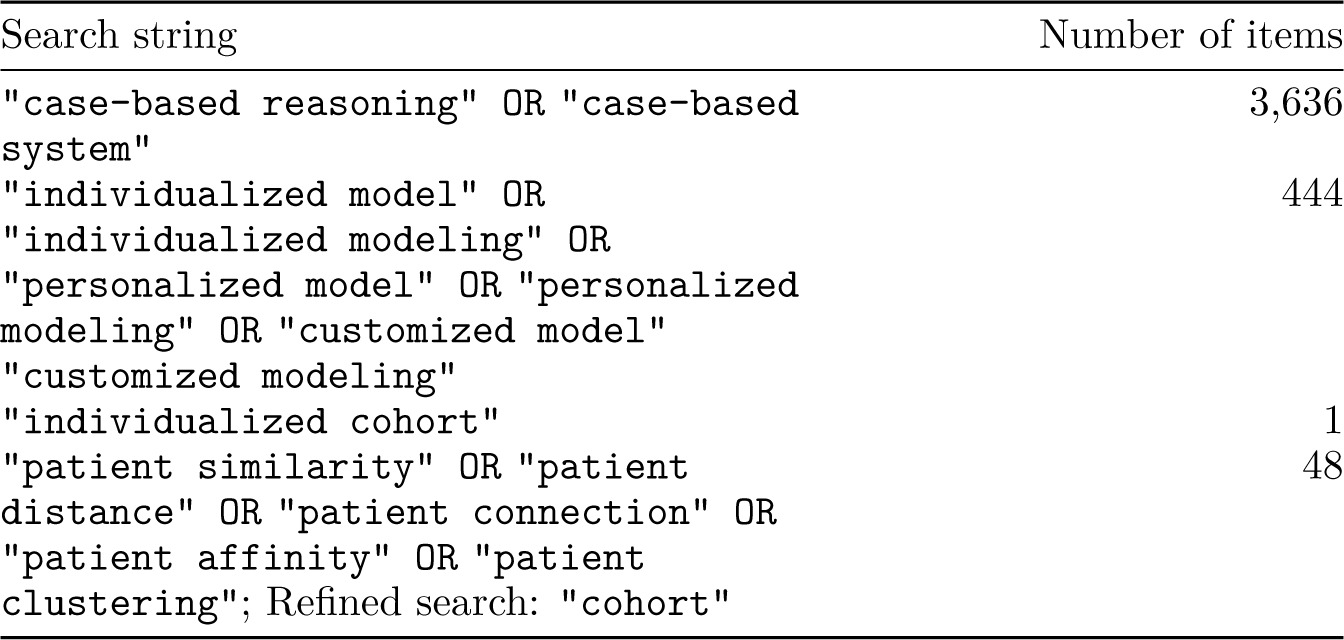

We found the results of the first search string to be predominantly irrelevant. To reduce review time, these were dropped. The last search was refined with an additional term following the initial disjunctive search. Our searches on Web of Science thus yielded 493 results.

When searching **Academic Search Premier**, we checked the option “Scholarly (Peer-Reviewed) Journals”, unchecked the option “Apply equivalent subjects”, and searched for several separate disjunctions and conjunctions of strings in the “TX All Text” field. We obtained the resulting citations via email in RIS format.

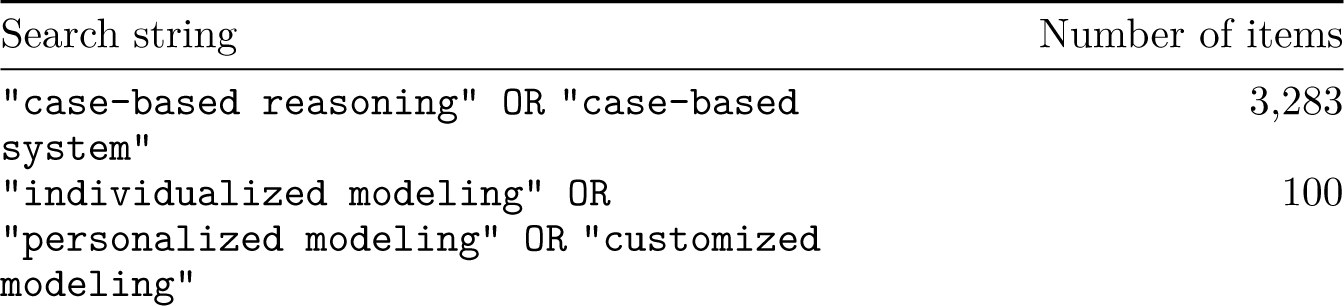
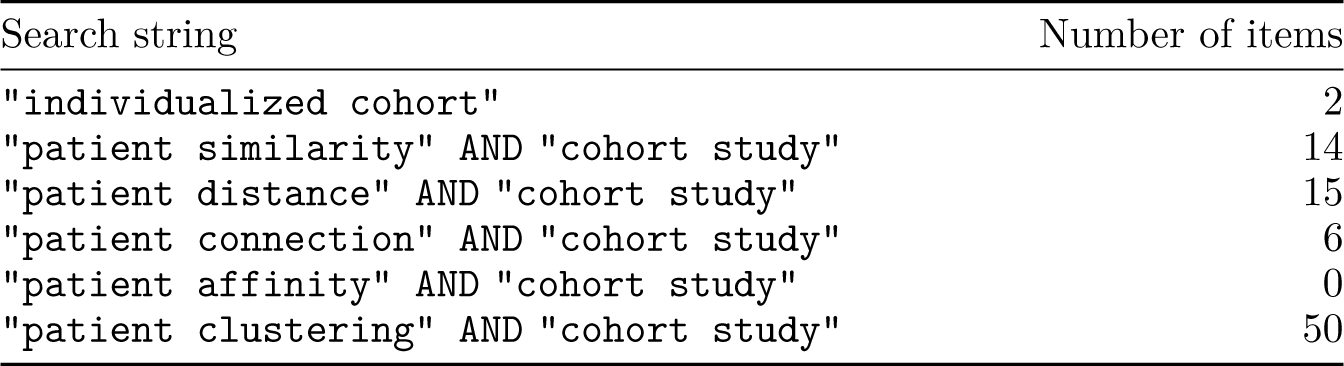

We found the results of the first search string to be predominantly irrelevant. To reduce review time, these were dropped. Our searches on Academic Search Premier thus yielded 187 results.

For **MathSciNet**, the portal to the *Mathematical Reviews* database, we used the same separate searches as for Web of Science, in some cases expanded to obtain more results. Those which yielded nonzero numbers of results are below:

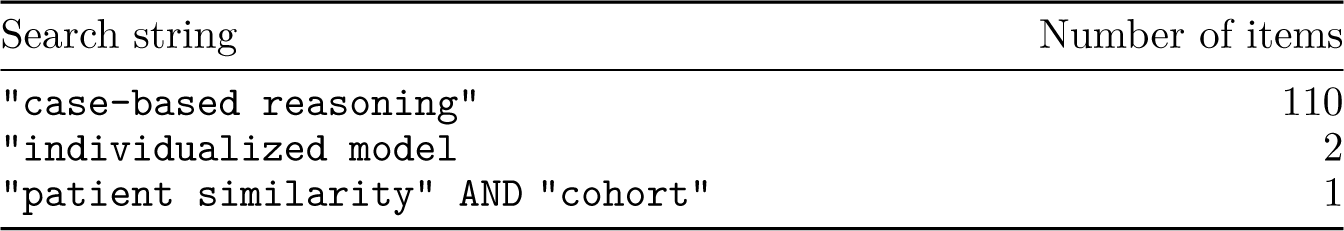

Our searches of *Mathematical Reviews* therefore yielded 113 results.

These totaled 1,422 sources from all platforms. We organized the full results in a public Zotero collection alongside the seed set and created a single folder for the 25 results reviewed in detail.

### Screening process

Most deduplication was done automatically in Covidence. As full-text review was done in Zotero, some additional duplicates were noticed and merged.

When abstracts were not obtained by search or by Covidence, we attempted to find them online using DOIs; when an abstract could not be found, screening was based on the title alone. We decided to screen titles and abstracts conservatively, rejecting only studies that were clearly outside the scope of our review. The reasons for rejection were four, as discussed in the main text: a. Exclude non clinical/non medical setting b. Must be in English c. Must be original study (not reviews, surveys, opinion, news) d. Exclude if search term clearly has different meaning than intended (Two examples of (d) are the use of the term “personalized model” to refer in some cases to parameterized models tuned to individual patient measurements and in others to patient-centered models of care.) Studies that passed title/abstract screen were exported to Zotero.

### Selection process

Some PDFs were obtained using Zotero from a university workstation, and the remaining were obtained through university library services. The review process is detailed in Section 2.4.

Our selection of relevant papers from the search corpus was based on the following inclusion/exclusion criteria:

- Uses labeled case-level (empirical) data set
- Defines a continuous-valued multivariate case similarity measure
- Uses the similarity measure to select cohorts for index cases from the corpus
- Fits statistical models to cohorts to make inferences about index cases

We decided after concluding full-text review to perform one round of citation-tracking, of citations within the Methods (or analogous) sections of the included entries.

### Update

Of the 25 studies included, 4 showed up in no database searches (but included from the seed set), 1 was obtained via reference tracking, and, of the remaining 20 found through the database searches, only 1 was not found in PubMed or Web of Science. That one was instead found in Mathematical Reviews. Since MR returns about the same volume of results as WoS and PM, we omitted it from the update.

We applied the search terms exactly as before but restricted the dates to 2021 July 19 (the beginning of the date range for our original searches) or later. We omitted the WoS search that returned impractically many results the first time around and was omitted then.

We ran the update search on 2024 January 24.

The PubMed update returned 97 results. The Web of Science refresher returned 272 + 0 + 26 = 298 results. Covidence removed duplicates, so that 348 studies were slated for screening.

Screening by title and abstract was done in two waves. First, PM excluded only on account of a study not being (a) clinical/medical, (b) English, and (c) original. Then, JCB then excluded on account of a study being (d) indicated by the intended meanings of our search terms.

### Analysis and synthesis

Because we could not predict the scope of methodological approaches we would encounter, we did not prepare specific analyses or syntheses *a priori*.

### Methodological elements

Table 6 and Figure 5 summarize the techniques and terminology used by the included studies, as referred to in the main text.

**Figure 5:**
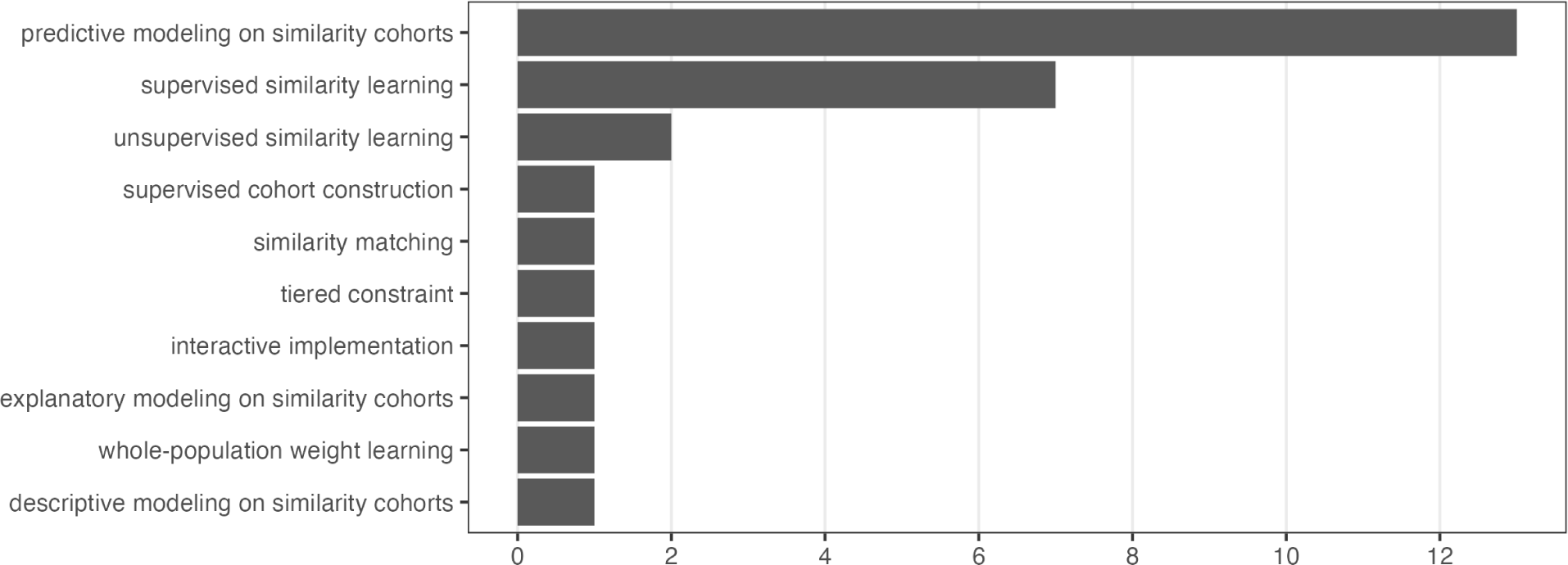
Frequency of recurring methodological elements across included studies.

### Performance evaluations and comparisons

Table 7 summarizes the evaluations and comparisons made of methods proposed in the included studies. In most cases, not all performance results are included; for the sake of concision, only what we judged to be the headline results, using the primary evaluation metrics, are reproduced here. The table contains no original results.

## 4.7 Data and code

Search results are publicly available in a Zotero group library. Data collected or encoded for included studies are publicly available in two Google Sheets. Code used to analyze these data and generate this manuscript is publicly available in a GitHub repository.

- Search results, included studies, and other bibliography: https://www.zotero.org/groups/5017571/imsr/
- Bibliographic and methodological properties of included studies: https://docs.google.com/spreadsheets/d/1tpWMhYH2pyRT55K7n2J2XFs-kEV_JTuCDmXzJ4BBgNo/
- Terminology and composite techniques of included studies: https://docs.google.com/spreadsheets/d/1xvDJwiLBoI2oz8fxHJ5MjNmiju_RAlK7RJv-wXe1DAs/
- Code used to conduct analyses and prepare the manuscript: https://github.com/corybrunson/imsr

## Footnotes

1 Similarity measures between more granular units of analysis, such as encounters and decision points, are often still referred to as patient similarity measures.

2 We note, in response to one reviewer’s comment, that for many if not most models used in ML, excepting GLMs, a predictor may play a different and more or less important role for some cases than others, as quantified by importance measures or model-agnostic explanations (Biecek and Burzykowski 2021; Molnar 2023). In this way, such models can also be viewed as “localized”. We recognize that the use of global and local explanations is standard. As another reviewer pointed out, the terminology of local and global is also used in the unrelated context of federated ML to distinguish models trained using data stored in a single storage device (local) from their aggregations (global) (Moshawrab et al. 2023; Brauneck et al. 2023). The prospects for federated architecture to more effectively implement similarity cohort–based models are intriguing, and would require some reconciliation of terms. At present, we feel that these methods are sufficiently disjoint to avoid confusion in practice, but we suggest the more specific term “similarity cohort–based models” when the term “local” is overloaded.

3 Drawing from Goyal, Lifshits, and Schütze (2008), we suggest the term “combinatorial CBR” for the then-conventional approach using similarity cohorts of fixed cardinality.

4 https://pubmed.ncbi.nlm.nih.gov/?term=(+"case-based+reasoning"+[All+Fields]+OR+"case-based+system"+[All+Fields]+)+OR+(+"individualized+modeling"+[All+Fields]+OR+"personalized+modeling"+[All+Fields]+OR+"customized+modeling"+[All+Fields]+)+OR+(+"individualized+cohort"+[All+Fields]+)+OR+(+(+"patient+similarity"+[All+Fields]+OR+"patient+distance"+[All+Fields]+OR+"patient+connection"+[All+Fields]+OR+"patient+affinity"+[All+Fields]+OR+"patient+clustering"+[All+Fields]+)+AND+(+"cohort+study"+[All+Fields]+)+)&sort=date

